# What will it take to achieve the End TB targets in South Africa? A mathematical modelling analysis

**DOI:** 10.64898/2026.04.23.26351599

**Authors:** Leigh F. Johnson, Mmamapudi Kubjane, Jeffrey W. Imai-Eaton, Lauren R. Brown, Lise Jamieson, Pren Naidoo, Gaurang Tanna, Gesine Meyer-Rath

## Abstract

**Background:** The WHO End TB strategy targets 80% and 90% reductions in TB incidence and mortality, respectively, between 2015 and 2030.

**Objective:** We assess which epidemiologic factors, including existing and new interventions, are most critical to reducing future TB in South Africa.

**Methods:** We adapted an existing mathematical model of TB and HIV in South Africa. Prior distributions were specified to represent uncertainty ranges for 27 model parameters that are highly uncertain and potentially important in driving future TB dynamics. Latin Hypercube Sampling was used to sample 1000 parameter combinations from these distributions, and the model was projected to 2040 for each. Partial rank correlation coefficients (PRCCs) were calculated to assess correlation between each parameter and average adult TB incidence and mortality rates over 2025-2040.

**Results:** Adult TB incidence and mortality rates in South Africa are projected to decline by 46% (95% CI: 17-69%) and 54% (95% CI: 21-84%) respectively by 2030, relative to 2015. The parameters most strongly associated with future TB incidence are the increase in microbiological testing in symptomatic individuals due to near-point-of-care/tongue swab (NPOC/TS) testing (PRCC=-0.67), reductions in social contact rates post-COVID (PRCC=-0.61), the probability of sputum testing in symptomatic individuals in the absence of NPOC/TS testing (PRCC=-0.39), and the efficacy of TB preventive therapy (PRCC=-0.35). TB mortality predictors are similar.

**Conclusions:** Increasing testing among people with TB symptoms, including through new NPOC/TS technologies, is likely to have the largest impact on progress towards End TB goals in South Africa, though attainment by 2030 is unlikely.

## Introduction

The End TB programme has set ambitious targets to reduce TB incidence by 80% and mortality by 90% globally, by 2030 (both relative to 2015 levels) [1]. Many new TB interventions have the potential to reduce TB incidence and mortality, including nutrition support [2], novel vaccines [3], targeted universal TB testing (TUTT) [4], community-based testing [5], computer-aided digital chest X-ray (dCXR) interpretation [6], tongue swab (TS) testing [7], near-point-of-care (NPOC) testing [8], and shorter drug regimens [9]. However, there is scepticism as to whether the End TB targets are achievable, given poor implementation of existing TB policies [10], especially in the wake of disruptions to TB services during the COVID-19 pandemic [11] and more recent cuts to funding of TB and HIV programmes [12]. There is also concern that the social determinants of TB (such as poverty and tobacco control) need to be addressed more systematically if TB incidence is to be reduced substantially [13, 14].

South Africa has one of the highest TB incidence rates in the world [15]. Dramatic increases in TB occurred during the late 1990s and early 2000s as a result of the rapidly expanding HIV epidemic, but TB incidence declined in the late 2000s following the successful introduction of a national antiretroviral treatment (ART) programme and intensified TB testing [16]. Since 2018, the number of microbiological diagnoses has stabilized at around 200 000 per annum, albeit with a brief dip during the COVID-19 pandemic [17, 18]. Future trends are highly uncertain. In addition to the uncertainty regarding the future implementation of interventions that form part of the TB programme, there is fundamental uncertainty regarding future social contact patterns, HIV risk behaviours and other factors, which are outside the control of the TB programme.

Mathematical models have an important role to play in quantifying these uncertainties. Using a modelling approach, we aim to assess which factors are most critical to reducing future TB incidence and mortality in South Africa, including factors that are changeable through policy and programme investments (such as current programme coverage and potential new interventions) and factors that might not be easily influenced but that are highly uncertain and potentially important drivers (such as social contact patterns, treatment engagement and efficacy of novel interventions). Our goal is to identify the most critical gaps in existing interventions, the new interventions that need to be prioritized, and the data gaps that require further research and monitoring, in order to inform the long-term strategy to reduce future TB incidence and mortality in South Africa.

## Methods

This analysis builds on the Thembisa model, an integrated TB, HIV and demographic model developed for South Africa [16, 17]. A detailed description of the model is provided elsewhere [17]; for this analysis, we fixed most parameters at the values estimated previously, when calibrating the model to TB data up to 2024 [17]. When projecting beyond 2024, we vary 27 parameters that are highly uncertain and potentially important in driving future TB dynamics. For each of the 27 parameters, a prior distribution is specified, representing the plausible range of uncertainty around the parameter of interest. These prior distributions are summarized in Table 1; a more detailed description of each parameter and the data sources on which the prior distributions are based is included in the supplementary materials.

**Table 1:** Prior distributions.

| Parameter | Baseline value<sup>a</sup> | Prior distribution | Prior mean, SD | 95% CI |
| --- | --- | --- | --- | --- |
| 1. TB contact rates and transmission |  |  |  |  |
| 1.1 Proportional preservation of behaviour change post-COVID | 0.50 | Beta (2.63, 2.63) | 0.50, 0.20 | 0.13-0.87 |
| 1.2 Reduction in TB contact rates due to AIPC in health facilities | 0 | Beta (2.15, 69.6) | 0.03, 0.02 | 0.00-0.08 |
| 2. HIV as a TB risk factor |  |  |  |  |
| 2.1 Relative rate of future ART initiation | 1 | Gamma (2.78, 2.78) | 1.00, 0.60 | 0.19-2.47 |
| 2.2 Change in condom use in future years | -0.09 | Normal (-0.09, 0.10) | -0.09, 0.10 | -0.29-0.11 |
| 2.3 Future rate of HIV testing, non-pregnant women aged 25 | 0.39 | Gamma (31.0, 79.6) | 0.39, 0.07 | 0.27-0.54 |
| 3. TB prevention |  |  |  |  |
| 3.1 Uptake of 3HP in PLHIV in 1<sup>st</sup> 6 months after ART start | 0.26 | Beta (3.87, 5.80) | 0.40, 0.15 | 0.13-0.71 |
| 3.2 Efficacy of 3HP in sterilizing infection | - | Uniform (0, 1) | 0.50, 0.29 | 0.03-0.98 |
| 3.3 Uptake of 3HP/TPT in household contacts | 0.08 | Beta (0.92, 2.77) | 0.25, 0.20 | 0.00-0.72 |
| 3.4 Time to introduction of nutritional support in household contacts, $T_n$ | - | Weibull (0.195, 0.55) | 33.1, 65.1 | 0-16<sup>+</sup><sup>b</sup> |
| Effectiveness of nutritional support in reducing TB incidence, $E_n$ | - | Beta (5.10, 7.98) | 0.39, 0.13 | 0.16-0.66 |
| 4. TB testing and diagnosis |  |  |  |  |
| 4.1 % of people seeking treatment for TB symptoms who receive laboratory-based sputum testing, $L$ | 0.10 | Beta (1.33, 2.27) | 0.125, 0.07 | 0.03-0.28 |
| Time to introduction of NPOC/TS testing in symptomatic adults, $T_t$ | - | Weibull (0.195, 0.55) | 33.1, 65.1 | 0-16<sup>+</sup><sup>b</sup> |
| % of people seeking treatment for TB symptoms who are tested if NPOC/TS testing is offered, $N$ | - | Beta (2.38, 2.29) | 0.51, 0.21 | 0.12-0.89 |
| 4.2 Proportion of eligible household contacts who are tested | 0.08 | Uniform (0, 0.6) | 0.30, 0.17 | 0.02-0.59 |
| 4.3 Average annual number of TUTT tests in people on ART | 0.22 | Gamma (1.44, 4.80) | 0.30, 0.25 | 0.02-0.95 |
| 4.4 Average annual number of TUTT tests in people with previous TB | - | Gamma (4.00, 8.00) | 0.50, 0.25 | 0.14-1.10 |
| 4.5 Time to introduction of door-to-door TB screening (symptoms), $T_d$ | - | Weibull (0.195, 0.55) | 33.1, 65.1 | 0-16<sup>+</sup><sup>b</sup> |
| Average annual screens per person by door-to-door screening, $S$ | - | Gamma (2.25, 12.5) | 0.18, 0.12 | 0.03-0.48 |
| 4.6 Proportion of door-to-door screened offered dCXR if asymptomatic, $P_x$ | - | Uniform (0, 1) | 0.50, 0.29 | 0.03-0.98 |
| 4.7 Proportion offered NPOC/TS test, in context of door-to-door screening, $P_t$ | - | Uniform (0, 1) | 0.50, 0.29 | 0.03-0.98 |
| 4.8 RR of culture testing if NAAT-negative, comparing future to current | 1 | Gamma (2.78, 2.78) | 1.00, 0.42 | 0.36-1.98 |
| 4.9 RR of TB screening in patients seeking treatment for TB symptoms, *R* | 11.1 | Gamma (2.86, 0.26) | 11.0, 6.5 | 2.2-26.9 |
| 5. TB treatment |  |  |  |  |
| 5.1 Initial LTFU after diagnosis in healthcare settings | 0.176 | Beta (10.0, 47.0) | 0.176, 0.05 | 0.09-0.28 |
| RR of initial LTFU after diagnosis in community settings | - | Beta (5.90, 2.70) | 0.68, 0.15 | 0.36-0.93 |
| 5.2 Time to introducing 4-month regimen for drug-sensitive TB | - | Weibull (0.195, 0.55) | 33.1, 65.1 | 0-16+<sup>b</sup> |
| 5.3 Relative rate of future treatment discontinuation | 1 | Gamma (2.97, 2.97) | 1.00, 0.58 | 0.20-2.42 |
| 5.4 OR for future cure rates, comparing future to current | 1 | Gamma (11.1, 11.1) | 1.00, 0.30 | 0.50-1.67 |

<sup>a</sup>No baseline value is specified for interventions that have not yet been introduced in South Africa. <sup>b</sup>A delay of more than 15 years means that the intervention is not introduced over the 15-year term of this analysis. 3HP = 3-month isoniazid and rifapentine regimen, AIPC = airborne infection prevention and control, ART = antiretroviral treatment, dCXR = digital chest X-ray, LTFU = loss to follow-up, NAAT = nucleic acid amplification test, NPOC/TS = near-point-of-care, with option of tongue swab/sputum specimens, OR = odds ratio, RR = relative rate, TPT = TB preventive therapy, TUTT = targeted universal testing for TB.

### Model description

Figure 1 provides a simplified representation of the model structure. People acquire *Mycobacterium tuberculosis* (*Mtb*) at a rate that depends on their daily number of close contacts, the prevalence of active TB, and probabilities of transmission per infectious contact. Two sources of uncertainty in future contact rates are considered. Firstly, some of the behaviour change observed during the COVID-19 pandemic (e.g. ‘self-isolation’ by people with respiratory infections, mask-wearing in congregate settings) may have continued in the post-COVID period. Secondly, efforts to improve airborne infection prevention and control (AIPC) in healthcare facilities could potentially reduce transmission.

**Figure 1:**
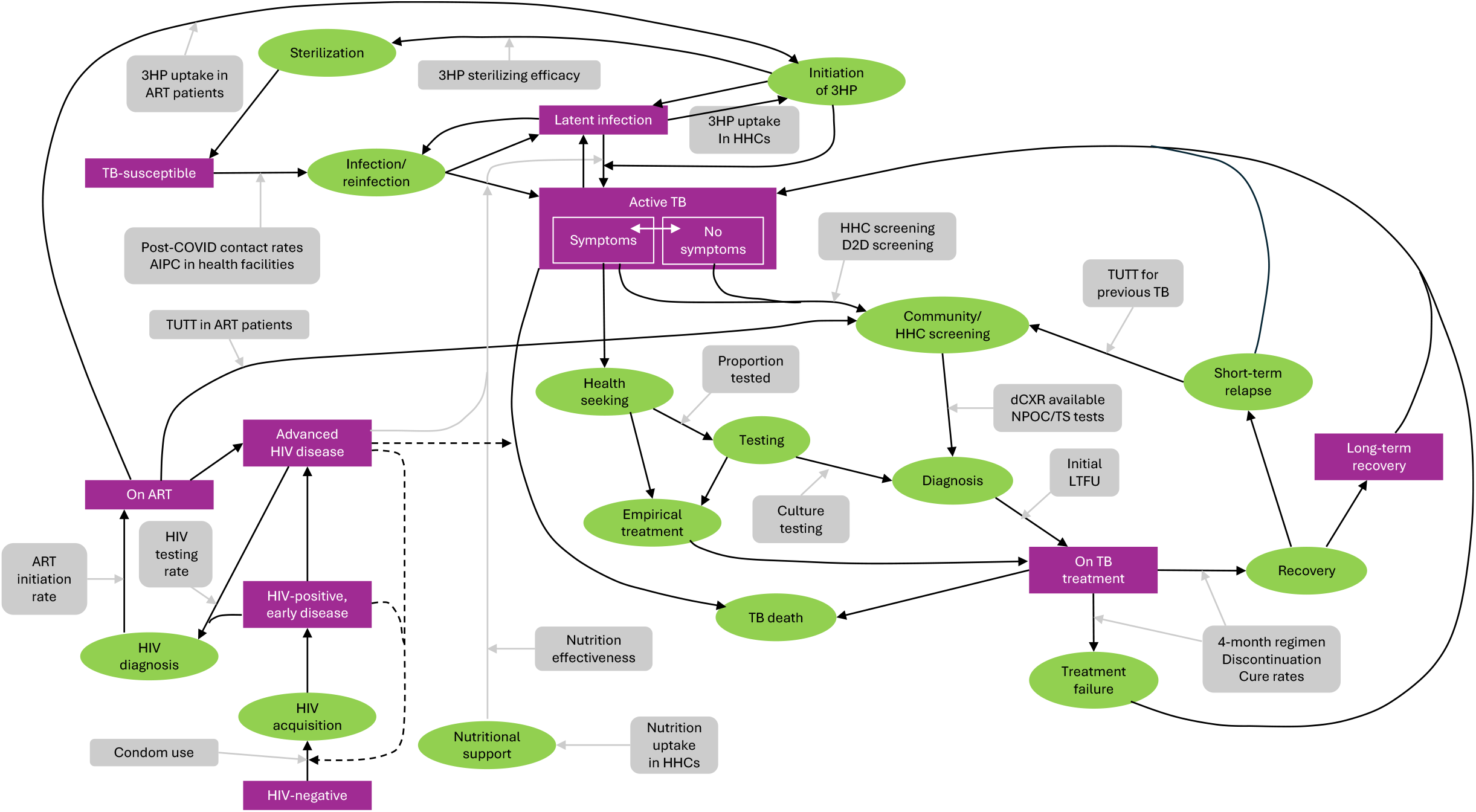
Simplified representation of TB model structure and the parameters included in the uncertainty analysis Purple rectangles represent disease states. Green ovals represent modelled events. Grey rounded rectangles represent model parameters that are varied in the uncertainty analysis and grey arrows represent the model processes that they influence. Solid black arrows represent transitions between disease states and events. Dashed black arrows represent effects of HIV prevalence/disease severity on HIV incidence and mortality. 3HP = 3-month isoniazid and rifapentine regimen, AIPC = airborne infection prevention and control, ART = antiretroviral treatment, D2D = door-to-door, dCXR = digital chest X-ray, HHC = household contact, LTFU = loss to follow-up, NPOC/TS = near-point-of-care, with option of tongue swab/sputum specimens, TPT = TB preventive therapy, TUTT = targeted universal testing for TB.

Upon acquiring *Mtb*, people either progress rapidly to active TB or remain latently infected, with the potential for active TB to develop years later, either as a result of ‘reactivation’ or reinfection with a new strain. The probability of rapid disease progression and the reactivation rate both depend on HIV status/disease stage. Three sources of uncertainty in future HIV programmes are considered: condom usage (which has historically been a major driver of reductions in HIV incidence [19], but which appears to be declining [20]), HIV testing, and the delay between HIV diagnosis and ART initiation (both of which could be negatively affected by recent US funding cuts [21]). The model also allows for the effect of four other risk factors on the TB disease progression/reactivation rates: under-nutrition, diabetes, smoking and binge drinking.

People with *Mtb* infection (but not active disease) can benefit from tuberculosis preventive therapy (TPT). South Africa has recently adopted a 3-month regimen of isoniazid and rifapentine (3HP) as the standard of care in TPT [22]. Although it is well established that 3HP reduces the incidence of active TB [23], it is less clear to what extent 3HP sterilizes *Mtb* infection; we therefore allow for uncertainty in this rate. South African guidelines recommend 3HP for people entering HIV care (almost all of whom are starting ART) and – more recently – all household contacts of people with TB [22]. The rates of future 3HP initiation in these two groups are allowed to vary in the uncertainty analysis. The model also allows for the possibility of a new policy of providing nutritional support to household contacts of people with TB, in order to reduce their TB risk [2]. Since this is not current South African policy, we consider uncertainty in both the timing of such a policy adoption and the effectiveness of nutrition support.

People with active TB who have TB symptoms are assumed to seek treatment at a rate that depends on their HIV status and sex (with higher rates for people with HIV and women). Although South African guidelines recommend nucleic acid amplification testing (NAAT) on sputum in all people with TB symptoms, only a minority of people with symptoms actually receive testing [24-26]; the future frequency of laboratory-based sputum testing in symptomatic individuals is allowed to vary in the uncertainty analysis. We also consider the possibility that microbiological testing rates might increase with the introduction of NPOC testing, which can be performed on TS or sputum specimens [8], in line with recent WHO guidance [27]. The yield on TS testing is assumed to be the same as for sputum testing [28]. Symptomatic individuals may also be tested for TB if they attend health facilities for other reasons, although this is assumed to be less likely, and the relative rate of testing in these individuals (compared to people seeking treatment for TB symptoms) is also allowed to vary in the uncertainty analysis. Culture testing is recommended for HIV-positive symptomatic individuals who test negative on a NAAT, and uncertainty in future rates of culture testing is also considered.

Community-based screening and TB testing in people without TB symptoms have historically been very limited, but TUTT has recently been introduced in South Africa. TUTT policy recommends routine annual testing, irrespective of symptoms, for three groups: people living with HIV (PLHIV), people who have been treated for TB within the last 24 months, and household contacts of TB patients. We consider uncertainty in TUTT rates in each of these three groups. Since door-to-door/community-based screening and testing is not current policy in South Africa, we allow for uncertainty in both the timing of such a policy adoption and the frequency of testing that would be achieved if the policy were adopted. The assumed standard for community-based testing is symptom screening (i.e. individuals are only tested if symptomatic), but this could be augmented through dCXR in people without symptoms or NPOC/TS testing; uncertainty in the adoption of both technologies is also considered. It is assumed that the uptake of community-based screening would double in the context of NPOC/TS testing [29, 30] (based on the lower cost, greater acceptability and quicker return of results [8, 31, 32]), but that the sensitivity of TS testing would be poorer than that of sputum testing (with relative sensitivities of 0.52 and 0.90 in asymptomatic and symptomatic TB respectively [7, 8]).

Following diagnosis, there may be initial loss to follow-up (LTFU), i.e. individuals do not start treatment despite a positive test result. Initiation of treatment is assumed to be less frequent if people are diagnosed outside of healthcare settings; both the future rate of initial LTFU in healthcare settings and the relative rate of treatment when diagnosed outside of healthcare settings are varied in the uncertainty analysis. People who start treatment may discontinue treatment prior to completion (at a rate that is varied in the uncertainty analysis) and have a probability of cure if completing treatment (also varied in the uncertainty analysis). Individuals who discontinue treatment have an increased risk of treatment failure, which implies continuation of active TB. The standard regimen for drug-sensitive TB has a 6-month duration, but 4-month regimens have been developed [9] and have been conditionally recommended by the WHO [33]. Since South Africa has not adopted the 4-month regimen, we allow for uncertainty regarding the timing of a potential policy change, assuming that such a regimen would lead to lower discontinuation but higher risk of treatment failure in people completing treatment [9].

### Selection of prior distributions

For each of the parameters varied in the uncertainty analysis, plausible ranges were assessed (lower and upper bounds), based on reviews of the literature and analysis of values estimated in the most recent calibration of the Thembisa model (where relevant). Prior distributions were then chosen such that the 2.5 and 97.5 percentiles of the distributions corresponded approximately to the lower and upper bounds respectively. In most cases, beta distributions were chosen to represent uncertainty in proportions (between 0 and 1), gamma distributions were chosen to represent uncertainty in positive-valued parameters (potentially greater than 1), and normal distributions were chosen otherwise. Weibull distributions were chosen to represent uncertainty in the time to specific policy changes; in all cases, the variable was specified in years from mid-2025, with a median of 10 years and a shape parameter of 0.55 (implying a 30% chance of the policy change occurring within 3 years and a 50% chance of the policy change occurring within 10 years).

### One-way sensitivity analyses

We defined the baseline scenario by setting all parameters to the prior means in Table 1, except in the case of the parameters relating to new interventions (nutritional support, NPOC/TS testing in symptomatic patients, door-to-door testing, and four-month TB treatment), which were excluded from the baseline scenario. For each parameter (except new intervention parameters), a one-way sensitivity analysis was conducted, varying the parameter between the lower and upper bounds (final column in Table 1) and calculating the percentage change in TB incidence/mortality over the 2025-2040 period, relative to that in the baseline scenario. For the new interventions, the sensitivity analysis considered immediate introduction (in 2025) and efficacy/uptake at the upper limit of the uncertainty range (only an upper bound on the intervention impact is displayed).

### Uncertainty analysis

From the 27 prior distributions specified in Table 1, a random sample of 1000 parameter combination was drawn, using Latin Hypercube Sampling [34]. For each parameter combination, the model was run to 2040, and trends in TB indicators were calculated. Partial rank correlation coefficients (PRCCs) were calculated to assess the correlation between each parameter and (a) the average TB incidence per 100 000 adults over the 2025-40 period and (b) the average TB mortality per 100 000 adults over the same period. (The PRCC differs from a regular correlation coefficient in that it allows for non-linearities in relationships and controls for variation in other parameters [34].) For the purpose of the PRCC calculations, some parameters were combined to create composite parameters that could be more easily interpreted (defined in Table 2), reducing the number of parameters in the PRCC analysis from 27 to 24.

**Table 2:** Composite parameters defined for the uncertainty analysis.

| Parameter | Calculation | Mean | SD |
| --- | --- | --- | --- |
| Reduction in TB risk in household contacts due to nutrition support | $E_n \frac{\max(0, 15 - T_n)}{2040 - 2025}$ | 0.165 | 0.182 |
| Increase in microbiological testing in symptomatic individuals due to introduction of NPOC/TS testing | $(N - L) \frac{\max(0, 15 - T_t)}{2040 - 2025}$ | 0.161 | 0.206 |
| Average annual probability of receiving symptom screening through door-to-door testing campaigns | $S(1 - P_t) \frac{\max(0, 15 - T_d)}{2040 - 2025}$ | 0.062 | 0.115 |
| Average annual probability of receiving dCXR screening through door-to-door testing campaigns, if asymptomatic | $SP_x(1 - P_t) \frac{\max(0, 15 - T_d)}{2040 - 2025}$ | 0.031 | 0.066 |
| Average annual probability of receiving NPOC/TS testing through door-to-door testing campaigns | $2SP_t \frac{\max(0, 15 - T_d)}{2040 - 2025}$ | 0.127 | 0.240 |
| Probability of microbiological testing in symptomatic individuals if seeking care for other conditions | $\left(L + (N - L) \frac{\max(0, 15 - T_t)}{2040 - 2025}\right) \frac{1}{R}$ | 0.042 | 0.058 |

## Results

Figure 2 shows model estimates of changes in TB incidence and mortality since 1990, in the baseline scenario (black lines, assuming no change to current programmes). TB incidence and mortality rates in South Africa peaked around 2004-5, then declined following the introduction of ART and increases in TB testing. In the absence of major changes to current programmes, adult TB incidence and mortality rates are expected to decline to 528 and 92 per 100 000 respectively by 2030, reflecting 36% and 40% reductions in TB incidence and mortality respectively, since 2015 – well below the End TB targets.

**Figure 2:**
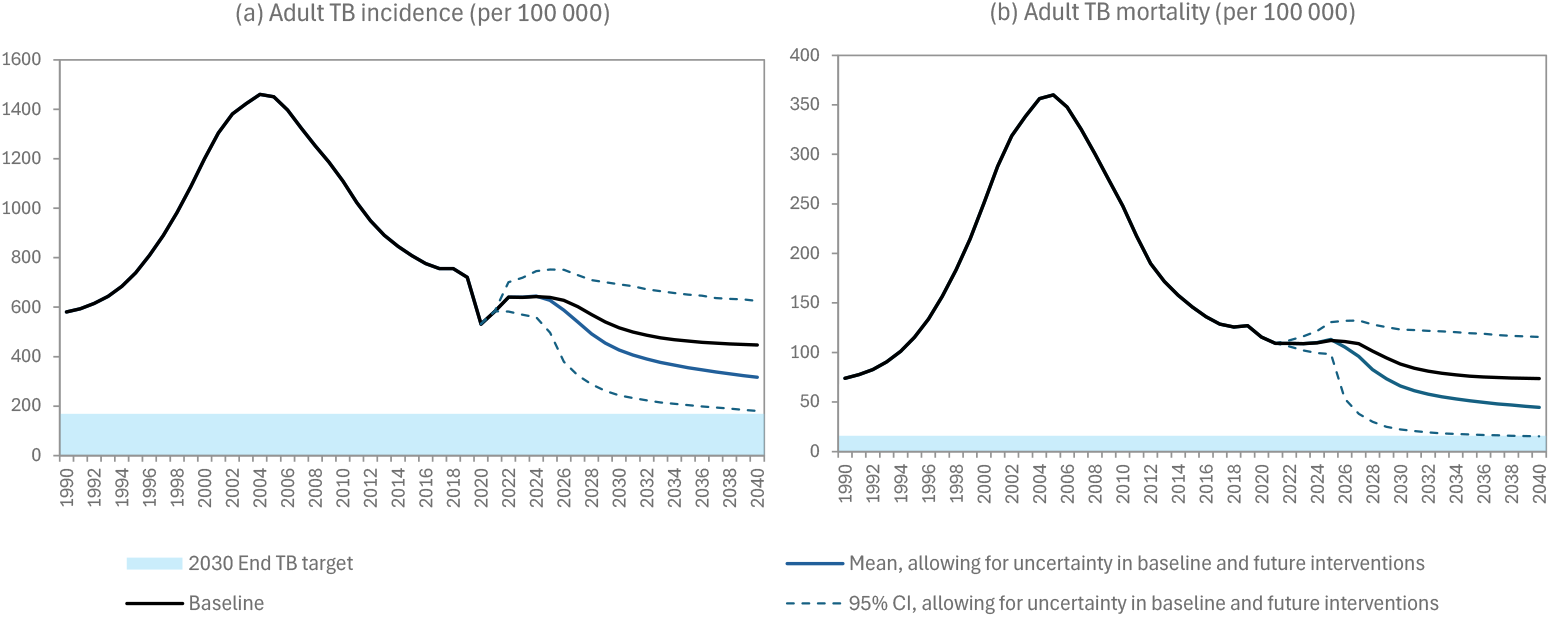
Trends in tuberculosis incidence (A) and mortality (B) Black line (baseline) represents expected future TB incidence and mortality in South Africa assuming continuation of current TB programmes and epidemiologic patterns, without introduction of new interventions. Blue line represents the mean over 1000 samples from the prior distributions of model parameters for epidemiologic parameters and future interventions (Table 1), including potential implementation of new interventions not currently implemented.

In one-way sensitivity analyses (Figure 3), the greatest reductions in TB would be achieved if NPOC/TS testing were implemented at 89% coverage in those seeking treatment for TB symptoms; this would achieve a 49% reduction in average TB incidence and a 67% reduction in average TB mortality over 2025-40. High rates of door-to-door testing with NPOC/TS could reduce TB incidence and mortality by 38% and 49% respectively, while combining high rates of door-to-door testing and dCXR screening in asymptomatic individuals could reduce TB incidence and mortality by 17% and 21% respectively. TB incidence could be reduced by 29% if levels of sputum testing in individuals seeking care for TB symptoms were increased to its upper bound, but would increase by 27% at the lower bound of future sputum testing coverage. Other influential parameters include the extent to which COVID-related behaviour changes are sustained, the future rate of ART initiation, and the combined uptake and efficacy of 3HP in people starting ART. Changes to the remaining parameters would not change future TB incidence or mortality by more than 10%.

**Figure 3:**
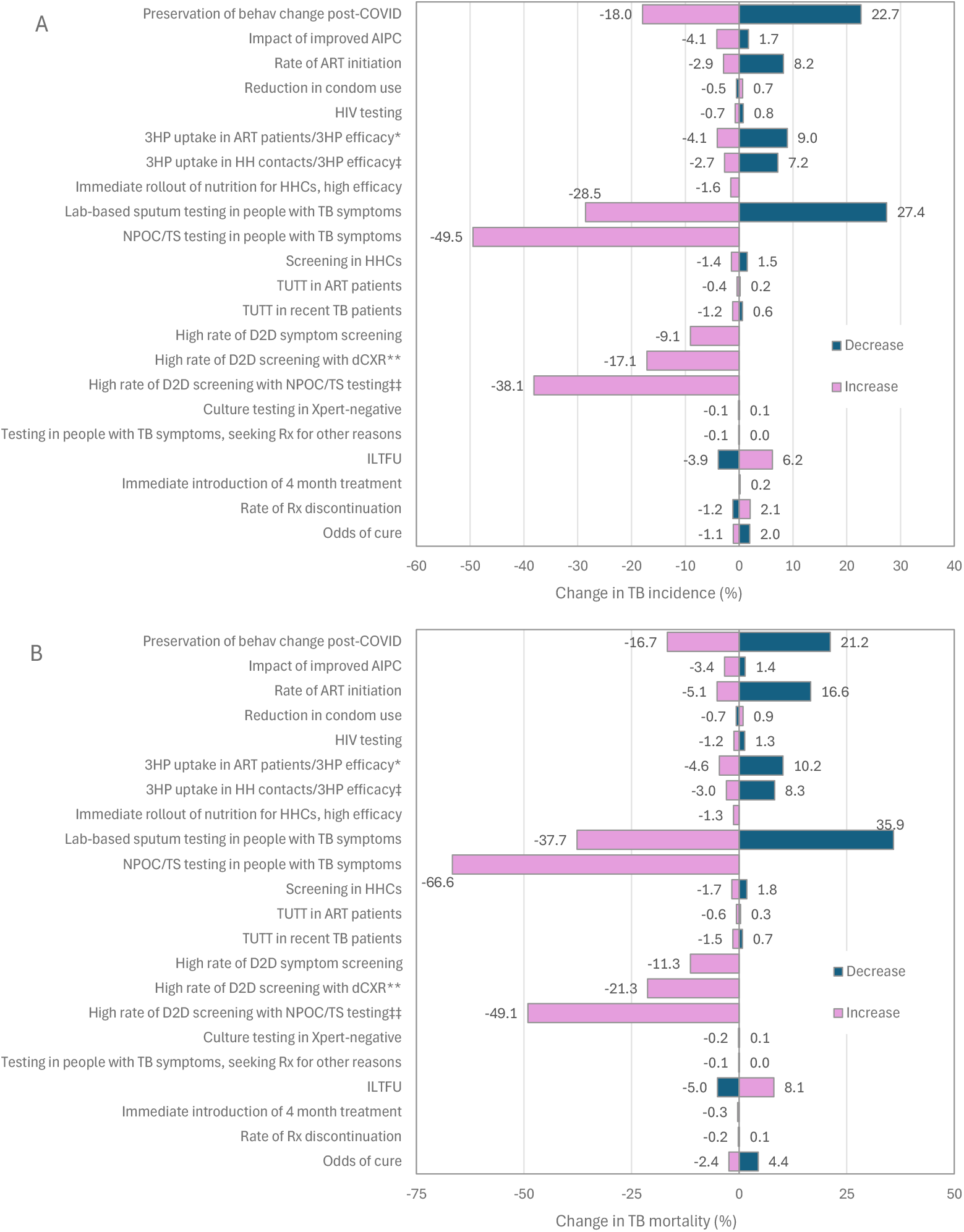
Percentage changes in average adult TB incidence (A) and mortality (B) over 2025-40 associated with increases and decreases to individual model parameters * Using upper limit of 3HP uptake in ART patients (parameter 3.1) and upper limit of 3HP efficacy (parameter 3.2). ‡ Using upper limit of 3HP uptake in household contacts (parameter 3.3) and upper limit of 3HP efficacy (parameter 3.2). ** Using upper limit of door-to-door testing frequency (parameter 4.5) and upper limit of proportion of those reached who are offered dCXR if asymptomatic (parameter 4.6). ‡‡ Using upper limit of door-to-door testing frequency (parameter 4.5) and upper limit of proportion of those reached who are offered tongue swab test if unable to produce sputum (parameter 4.7). 3HP = 3-month isoniazid and rifapentine regimen, AIPC = airborne infection prevention and control, ART = antiretroviral treatment, D2D = door-to-door, dCXR = digital chest X-ray, HHC = household contact, LTFU = loss to follow-up, NPOC/TS = near-point-of-care, with option of tongue swab/sputum specimens, TUTT = targeted universal testing for TB.

Figure 2 shows the model projections to 2040 in the uncertainty analysis (blue lines, allowing for uncertainty in baseline parameters and uncertainty in potential new interventions). The adult TB incidence rate in South Africa is projected to decline by 46% by 2030, relative to 2015 levels (95% confidence interval [CI]: 17-69%), and the adult TB mortality rate is projected to decline by 54% (95% CI: 21-84%) over the same period. By 2030, a more than 70% reduction in incidence is achieved in 1.8% of scenarios, and a more than 80% reduction in mortality is achieved in 10.6% of scenarios. Greater reductions are forecast when projecting the decline from 2015 to 2040 for TB incidence (62%, 95% CI: 25-78%) and TB mortality (71%, 95% CI: 26-90%).

Figure 4 shows the correlations (PRCCs) between each of the 24 parameters and the average TB incidence rate (panel A) and average TB mortality rate (panel B). The parameters most strongly associated with future TB incidence are the increase in microbiological testing in symptomatic individuals through NPOC/TS testing (PRCC=-0.67), reductions in social contact rates post-COVID (PRCC=-0.61), the probability of sputum testing in symptomatic individuals in the absence of NPOC/TS testing (PRCC=-0.39), the efficacy of 3HP in sterilizing infection (PRCC=-0.35), and the probability of testing in people with TB symptoms who are seeking care for other conditions (PRCC=-0.30). Introducing door-to-door/community-based symptom screening and testing could significantly reduce TB incidence, particularly if combined with NPOC/TS testing. TUTT is not generally expected to significantly reduce TB incidence.

**Figure 4:**
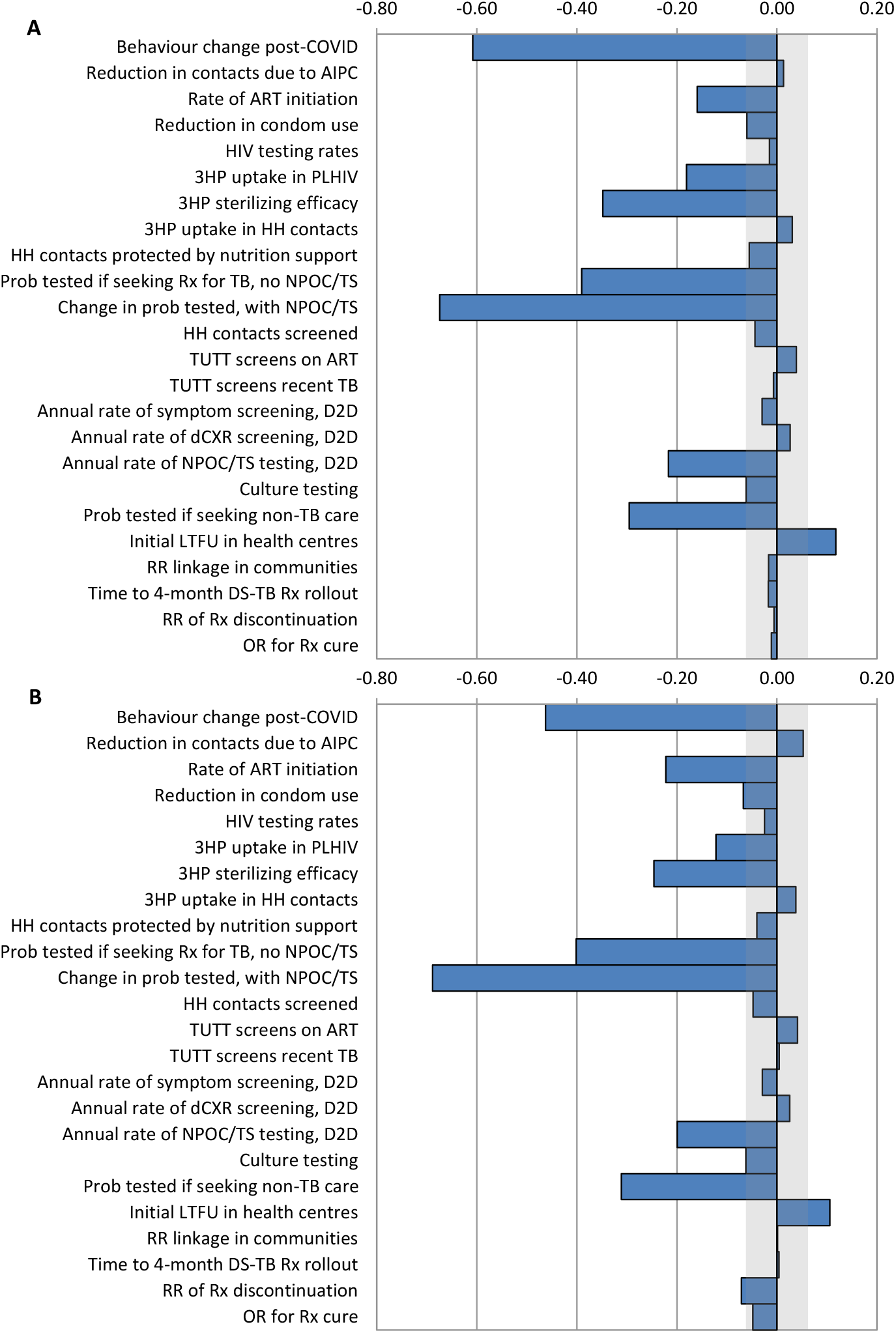
Partial rank correlation coefficients between 24 parameters and average adult TB incidence rates (A) and mortality rates (B) in South Africa, over 2025-2040 The shaded grey area represents the range in which correlation coefficients that are not significantly different from zero. 3HP = 3 months of isoniazid and rifapentine (TB prevention), AIPC = airborne infection prevention and control, ART = antiretroviral treatment, D2D = door-to-door (community-based testing), dCXR = digital chest X-ray, DS-TB Rx = drug-sensitive TB treatment, HH = household, LTFU = loss to follow-up, NPOC/TS = near-point-of-care, with option of tongue swab/sputum specimens, PHC = primary healthcare facility, PLHIV = people living with HIV, RR = relative rate, TUTT = targeted universal TB testing.

The factors that most influence TB mortality are similar to those driving TB incidence, with the most significant determinants of TB mortality again being the increase in microbiological testing in symptomatic individuals due to NPOC/TS testing (PRCC=-0.69), the reduction in social contacts post-COVID (PRCC=-0.46), the probability of sputum testing in people seeking treatment for TB symptoms in the absence of NPOC/TS testing (PRCC=-0.40), the probability of testing in people with TB symptoms who are seeking care for other conditions (PRCC=-0.31), the efficacy of 3HP in sterilizing infection (PRCC=-0.25), and the rate of ART initiation (PRCC=-0.22).

## Discussion

In the absence of future new technologies it is unlikely that South Africa will meet the 2030 End TB targets, although it is possible to achieve reductions close to the targets, particularly if NPOC/TS testing proves to be transformative in increasing the uptake of testing. Previous modelling studies have also suggested that South Africa and other countries in sub-Saharan Africa are unlikely to meet the 2030 target in the absence of a TB vaccine [35-37].

We found that the rate of testing in people seeking treatment for TB symptoms is the most important driver of future TB incidence and mortality. This finding is not surprising when considering how low historic rates of TB testing have been. In studies of patients seeking treatment for TB symptoms in South African health facilities, the proportion who received a TB test has been highly variable, between 3% and 84% [24-26, 38], with a median of only 30%. Similarly low rates have been reported in other countries with high TB burdens [10]. These low rates of testing reflect the non-specificity of TB symptoms and healthcare workers’ concerns about the cost and time required to collect sputum specimens and send them to a central laboratory for NAAT [39]. In one South African study, for example, 36% of all clinic attenders had symptoms suggestive of TB [40] – conducting TB testing in such a large fraction of primary care attendees may well be infeasible [10]. Achieving high levels of testing in people with TB symptoms could require levels of laboratory testing beyond current testing capacity. However, new point-of-care tests performed on tongue swabs or sputum present an important opportunity to increase levels of TB testing in symptomatic individuals at reduced cost, and without placing additional strain on laboratories [8, 31]. Regardless of the testing platform, there is a need to strengthen adherence to existing guidelines for systematic testing of all patients with symptoms suggestive of TB [41]. There is also a need for an increased healthcare workforce dedicated to TB diagnosis, and better monitoring systems to identify where and why symptomatic patients are not being tested.

Another major determinant of future TB incidence is recent changes in social contact rates – this refers both to general rates of social mixing and measures taken by people with TB symptoms to reduce their risk of transmission to others. Although there is strong evidence of reductions in social contact rates during COVID-19 in South Africa [42-44], it is not clear to what extent risk behaviours have reverted to pre-COVID levels. It might be assumed that the heightened awareness of the importance of mask wearing and self-isolation by people with respiratory infections during the COVID-19 pandemic would have had an enduring effect post-pandemic, and mask wearing is still evident in South Africa today, albeit uncommon. The lack of reliable, current data makes it difficult to determine this parameter reliably. However, our results suggest that there may be continued value in public awareness campaigns to promote mask wearing and social isolation by people with respiratory symptoms. There may also be need for targeted promotion of mask wearing in settings such as healthcare facilities and public transport, and the National TB Plan calls for mandatory mask wearing in all health facilities [45].

Previous work suggests that isoniazid preventive therapy (IPT) targeted to PLHIV has had only a very modest impact on TB incidence in South Africa at a population level [16, 46]. However, IPT provides only short-term protection against TB – protection is rapidly lost after people discontinue IPT in African settings [47-49]. 3HP, in contrast, sterilizes a proportion of TB infections [50], ensuring a more long-term reduction in TB risk [23, 49, 51]. The exact magnitude of this proportion is difficult to estimate, but our results are consistent with other modelling studies, which suggest that the population-level impact of 3HP is highly dependent on the fraction sterilized – more so than any other dimension of efficacy [52]. Although the replacement of IPT by 3HP was announced in South Africa in 2021, coverage of 3HP remains low, and needs to be increased, especially in PLHIV.

Community-based strategies are likely to have relatively little impact on TB incidence if testing is conducted only in symptomatic individuals. Previous reviews have noted that active case finding (ACF) tends to detect less severe TB (more smear-negative TB and less severe cavitation), and the overall impact of ACF at a population level is modest [53]. However, a recent trial in Vietnam showed that with very high levels of community-based testing, including in asymptomatic individuals, it is possible to achieve significant short-term reductions in TB incidence [5], and consistent with this, our results suggest substantial impacts are possible when community screening interventions include either dCXR screening or NPOC/TS testing in asymptomatic individuals. However, the feasibility of sustaining such intense screening over the long term is debatable. Another concern is that although TS testing has good sensitivity in symptomatic individuals, its sensitivity in individuals with asymptomatic TB appears poor [7]. We assume that this lower sensitivity is largely offset by higher acceptability and greater ease of providing TS specimens, but much uncertainty remains around the likely performance of TS testing in community settings.

We also find that TUTT generally is expected to have relatively little population-level impact. In the first TUTT trial in South Africa, the intervention had only a modest effect: a 17% increase in the annual number of TB cases treated [4]. Our model assumes that these additional asymptomatic individuals have relatively low infectiousness and mortality risk when compared to those with symptomatic TB, which partly explains why there is not a larger impact on TB incidence and mortality. We also find in our one-way sensitivity analysis that TUTT in household contacts and recently-treated TB patients has a greater impact than TUTT in PLHIV. In the TUTT trial, the test positivity rates were highest in people with prior TB and lowest in PLHIV [54]. We assume a high rate of relapse during the first 6 months after treatment completion [55], which might explain why we estimate a larger benefit from screening people who have recently had TB than for PLHIV.

Our results suggest that rates of ART initiation and 3HP uptake in ART patients are likely to be important determinants of future TB incidence and mortality, consistent with previous modelling studies showing that HIV and ART have strongly determined TB incidence in South Africa [16, 56-58]. However, we find that the benefits from increased TB testing in PLHIV are questionable. It should be noted that PLHIV contribute relatively little to *Mtb* transmission [59-61], i.e. the secondary benefits of improved detection and treatment of TB in PLHIV (in terms of reducing *Mtb* transmission) are likely to be modest. High levels of TB testing in PLHIV have already been achieved in South Africa [62], while rates of TB testing in the HIV-negative population are relatively low, which explains why the marginal gains from further increasing TB testing in PLHIV are estimated to be small.

Consistent with more detailed modelling of nutritional support by McQuaid *et al* [63], we estimated that this intervention will have relatively little impact on future TB incidence and mortality. Although the intervention is effective in reducing TB risk, we assume that (as in a recent trial [2]), it would only be offered to household contacts of TB patients. In the South African context, within-household transmission is estimated to account for less than 20% of *Mtb* transmission [64, 65], and thus an intervention targeting household contacts might be expected to have only a modest impact. This also explains why 3HP in household contacts and TUTT in household contacts is predicted not to have a substantial impact at a population level. Other modelling studies also suggest tracing of household contacts would have only modest impacts on TB incidence at a population level [66].

This study is not a cost-effectiveness analysis. The interventions that we have identified as being potentially impactful are not necessarily interventions that would be affordable or cost-effective. Conversely, interventions that do not have a substantial impact relative to other interventions may nevertheless be cost-effective. Recent analysis of the relative cost-effectiveness of TB interventions in South Africa suggests that increasing rates of NAAT testing in people with TB symptoms attending health facilities could be cost-saving [67]. Our study also does not consider the potential impact of TB vaccines, as our intention is to focus on currently available TB prevention and treatment strategies. Previous modelling studies suggest TB vaccination could have a dramatic impact in South Africa [68], but currently it is unclear when the first vaccines will be distributed and how effective they will be. Further limitations are that our model does not include paediatric TB or differences between drug-sensitive and drug-resistant TB, though these are likely to be minor drivers of *Mtb* transmission in South Africa. We have also not considered uncertainty related to risk factors other than HIV (smoking, alcohol, under-nutrition and diabetes), due to difficulties in quantifying the uncertainty around the future prevalence of these risk factors.

Although there is much reason to be hopeful about new TB technologies, achieving effective implementation of previously established interventions should not be forgotten in the push towards the End TB targets. South Africa’s TB strategic plan for 2023-28 has placed emphasis on new interventions such as community-based testing and novel diagnostics, but is largely silent on the poor implementation of previous guidelines [45]. Chakaya *et al* [11] note that the challenge in achieving significant reduction in TB “seems to be sub-optimal application of effective interventions” rather than a lack of effective interventions. Our study argues for striking a balance between adopting new interventions and addressing major gaps in the implementation of already-established interventions, with current gaps in testing patients with symptomatic TB and 3HP being areas of particular concern.

## Data Availability

This is a mathematical modelling study, which focuses on potential future scenarios. Although data have been used to calibrate the model, the calibration is not the focus of this paper, and is described in previous publications.

