## Supplementary materials for "What will it take to achieve the End TB targets in South Africa? A mathematical modelling analysis"

Prior distributions

This document describes the model parameters that are subject to the most future uncertainty, reviews data on plausible values for these parameters and describes the prior distributions assigned to represent uncertainty in these parameters.

### **1. TB contact rates and transmission**

#### **1.1 Change in contact rates post-COVID**

The 2023-2028 National Strategic Plan aims to reduce TB transmission in “high-risk indoor places where people congregate”, for example schools and churches, through community education and improving natural ventilation [1].

Our model incorporates changes in social contact rates in response to COVID-19, based on evidence of reductions in social contact rates in the early stages of the pandemic [2-4]. However, it is unclear to what extent contact rates have reverted to normal post-COVID, and it is also unclear whether the high-level goals in the NSP are likely to lead to real changes in social contact rates. One might assume that increased awareness of the importance of ‘self-isolation’ when experiencing symptoms of respiratory infection would imply a sustained reduction in rates of infectious contact, and increased normalization of mask wearing (especially in crowded spaces) may have also contributed to a sustained reduction in transmission risks. To represent the uncertainty around the contact rates after mid-2022, we multiply the pre-2020 contact rates by a factor of (1 – 0.4*R*), where 0.4 is the reduction that was assumed for the 2021-22 period and *R* is the post-2022 adjustment factor. We represent the uncertainty in *R* using a beta prior distribution with a mean of 0.5 and standard deviation of 0.2. This means that on average, contact rates in the post-2022 period are 20% lower than those pre-COVID, consistent with the current Thembisa assumptions [5].

#### **1.2 Infection control in health facilities**

One of the stated objectives of the 2023-2028 National Strategic Plan is to improve airborne infection prevention and control (AIPC) in health facilities, for example through improving access to face masks, and ‘decongesting’ health facilities by improving community-based distribution of drugs to patients with chronic conditions [1]. The TB Strategic Plan for 2023-28 further states the objective of rolling out ultraviolet germicidal irradiation (UVGI) in all clinics, and notes that this already occurred in Free State province during the COVID-19 pandemic [6]. Banholzer *et al* [7] estimate that in a Cape Town clinic almost all TB transmission was eliminated during the COVID-19 pandemic as a result of measures put in place to counter the spread of SARS-CoV-2, including mandatory face masks, physical distancing, decongestion and improved ventilation. However, it is not clear to what extent these measures have been sustained post-COVID, or how widely they have been adopted across South Africa.

Few studies have attempted to estimate the fraction of TB transmission that occurs in healthcare settings, in the African context. A recent study from Botswana estimated that 5% of transmission occurred in healthcare facilities, and noted that this was likely to be an upper bound [8]. A model of TB transmission in rural KwaZulu-Natal similarly estimated that 4.9% of transmission occurred in health facilities, and further estimated that improvements in AIPC in health facilities could reduce total TB incidence in the population, over 10 years, by 3.4-8.0% [9]. Based on these studies, we represent the proportional reduction in TB contacts due to improved AIPC using a beta distribution, with a mean of 3% and standard deviation of 2%. We chose a relatively low mean because of uncertainty around the extent to which improved AIPC will be introduced in South Africa. The 2.5 and 97.5 percentiles of this distribution are 0.4% and 8.0% respectively, and the upper bound therefore corresponds to the reduction that might optimistically be expected based on the KwaZulu-Natal modelling study.

### **2. HIV as a TB risk factor**

#### **2.1 Future delays in ART initiation**

To reflect uncertainty regarding future ART coverage, we allowed for uncertainty in the average delay between diagnosis and ART initiation (in those people who do not initiate ART immediately after diagnosis). In version 4.7 of the Thembisa model, for example, the assumed average delay between diagnosis and ART initiation (in women who did not start ART in the first month after diagnosis) was 14 months [10], equivalent to a monthly ART initiation rate 0.071 (1/14). However, this rate was reduced to 0.013 for the purpose of assessing a “minimum” impact that a PEPFAR funding withdrawal might have in South Africa, if there were no move by the South African Department of Health to take over the ART programme support provided by PEPFAR [11]. Somewhat antithetically, the Department of Health recently announced a plan to increase the total number of people on ART in South Africa by 1.1 million. In helping the Department of Health to forecast the like impact of such an ambitious target, we increased the ART initiation rate in women to 0.14 and assumed reductions in rates of ART interruption (Lise Jamieson, personal communication). It is worth noting that although the Thembisa model allows for people to cycle on and off ART, the model does not allow for these ART interruptions to affect TB incidence and mortality, which is why we focus here on changes to the rate at which people initiate ART after diagnosis (and not rates of ART interruption or re-initiation).

We apply a multiplier to the default future treatment initiation rates for men and women, and we represent the uncertainty in this multiplier using a gamma prior distribution with a mean of 1 and standard deviation of 0.6. This prior distribution has 2.5 and 97.5 percentiles of 0.19 and 2.47 respectively; the lower limit is thus roughly in line with the ratio assumed when modelling the potential effect of PEPFAR withdrawal (0.013/0.071 = 0.18), while the upper limit is roughly in line with the ratio assumed when modelling the achievement of the 1.1 million target (0.14/0.071 = 1.97, though other model changes are needed to achieve the full 1.1 million increase). Figure S1(d) shows that with these uncertainty ranges, we obtain reasonably wide ranges of uncertainty in future ART coverage; by 2040 the 2.5 and 97.5 percentiles of the distribution of ART coverage estimates are 72% and 93% respectively.

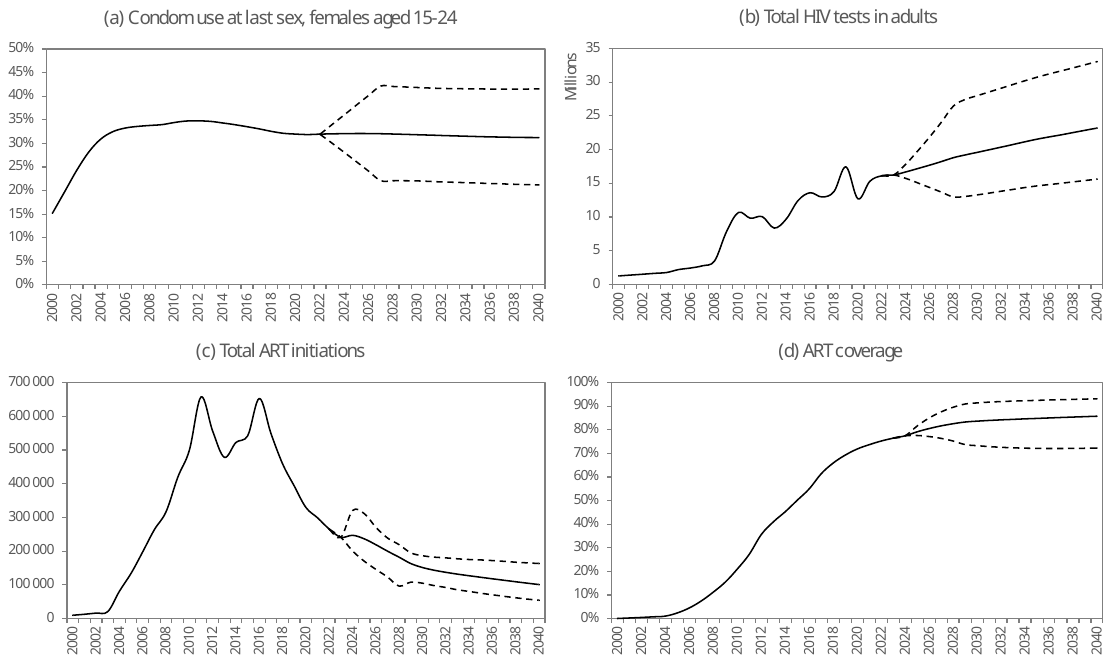

Figure S1: Projections of uncertainty in future HIV programme indicators

Dashed lines represent the 2.5 and 97.5 percentiles generated from a sample of 1000 HIV parameter combinations, drawn from the prior distributions described in sections 2.1, 2.2 and 2.3. The solid line represents the average of the 1000 results.

#### **2.2 Future trends in condom use**

In the Thembisa model, we make provision for potential future changes in condom use through the $k_{2}$ parameter, described more fully in section 2.8.1 of the Thembisa HIV model report [10]. This parameter was previously set to 0.016 based on fitting the model to historic data on self-reported condom use. This suggests a slight positive trend in condom use in recent years. However, this was based on data up to 2017. In the most recent version of Thembisa (version 4.8), we refitted the model after updating it to include condom use data from the 2022 South African National HIV Prevalence, Incidence, Behaviour and Communication survey [12], and the revised estimated of the $k_{2}$ parameter was -0.09. The lower-than-expected level of condom use in the survey coincides with declining levels of condom distribution in South Africa; for example, male condom distribution coverage is estimated to have declined from 36.8% in 2018 to 18.9% in 2022 [13]. However, it is difficult to infer true levels of condom use from these data, as there is estimated to be substantial condom wastage [14]. In addition, the self-reported condom usage data from the most recent 2022 survey have many uncertainties attached to them (including potential social desirability bias and sampling uncertainty reflected in the wide 95% confidence intervals). We therefore represent the uncertainty in the reduction in condom use in recent years using a normal distribution with a mean of -0.09 (the same as the Thembisa 4.8 model estimate) and a standard deviation of 0.10. Figure S1(a) shows that with these uncertainty ranges, we obtain reasonably wide ranges of uncertainty in future condom use; by 2030 the 2.5 and 97.5 percentiles of the distribution of condom usage rates in females aged 15-24 are 22% and 42% respectively.

#### **2.3 Future levels of HIV testing**

Levels of HIV testing in South Africa have been relatively stable in recent years, although there was a dip in levels of HIV testing during 2020-21 as a result of COVID-19 disruptions. In the most recent version of the Thembisa model (version 4.8), the annual rate of HIV testing in non-pregnant women aged 25, who have not previously been tested, is estimated to have been 0.39 on average between mid-2019 and mid-2024 [15]. Although it is not anticipated that this rate would change substantially in the near future, levels of HIV testing in several other African countries have dropped dramatically in recent years, driven partly by reductions in PEPFAR support for general HIV testing programmes [16]. South Africa’s HIV testing programme is less reliant on donor funding than that of other countries in the region. Nevertheless, there is the danger that levels of HIV testing could be scaled back significantly, especially in light of recent public health sector funding cuts and PEPFAR funding cuts. In a worst-case scenario it was recently assumed that HIV testing rates in South Africa might drop by as much as 30% due to the US funding cuts [11].

On the other hand, there is the possibility that levels of HIV testing may increase in future, especially with the increasing adoption of self-testing kits. Although numbers of self-testing kits distributed in the public sector remain low, self-testing kits are increasingly available in pharmacies, and there could be significant growth in private demand for HIV self-testing.

To represent the uncertainty regarding future HIV testing trends, we specify a prior distribution around the rate of HIV testing in non-pregnant women aged 25 who have not tested previously. (The multipliers that determine relative rates of testing in men and at different ages are held constant, and therefore changes to this parameter will also affect numbers of tests performed in men and older women.) We use a gamma prior distribution with a mean of 0.39 and a standard deviation of 0.07; this distribution has 2.5 and 97.5 percentiles of 0.27 and 0.54 respectively. The prior mean has thus been chosen to be consistent with the recent levels of HIV testing in South Africa, while the standard deviation was chosen so that the 2.5 percentile would be 30% lower than the current baseline (i.e. corresponding to the anticipated ‘worst case’ scenario for the impact of the US funding cuts). Figure S1(b) shows that with these uncertainty ranges, we obtain reasonably wide ranges of uncertainty in future levels of HIV testing. On average, the total number of HIV tests is predicted to increase as a result of population growth.

### **3. TB prevention**

#### **3.1 Uptake of 3HP in PLHIV starting ART**

In a meta-analysis of studies that assessed the proportion of people living with HIV (PLHIV) who started TPT [17], the average global estimate was 41% in studies in which there was no required testing for TB infection prior to TPT (as is current policy in South Africa). However, only 44% of PLHIV in these studies were on ART, and it is not clear how representative this estimate is of people starting ART.

In the most recently calibrated Thembisa TB model [5], we estimated that monthly rates of TPT initiation during the first 6 months of ART, in adults starting ART with CD4 counts of <200 cells/μl were between 0.05 and 0.10 over the 2010-2024 period, equivalent to cumulative probabilities of initiating TPT, over the first 6 months of ART, of 26-45% (average of 37%). Corresponding estimates from other South African studies are quite variable. In a small Cape Town study, only 17% of ART patients had ever initiated IPT [18], and similarly low proportions were found in two clinics in Gauteng (7% and 26%) [19]. In a KwaZulu-Natal study, the proportion was 48% and 71% in two different clinics [20]. In a 2011 national sample of health facilities, approximately 33% of people newly diagnosed with HIV initiated TPT, although it was not clear whether this proportion differed according to whether patients started ART [21].

South African studies have noted several barriers to TPT initiation, including non-availability of drugs, lack of healthcare worker awareness/training, concerns about potential drug resistance (and related to this, concern that standard symptoms screens are insufficient to exclude active TB), TPT being perceived as a ‘low priority’ in the context of over-burdened health services, and socio-economic factors that prevent patients from accessing health services [22, 23]. Many of these factors could be addressed through future interventions, especially if 3HP is promoted as being an easier and more effective approach to TB prevention than the previously used IPT. However, programme data suggest a downward trend in levels of TPT provision in recent years, and there is a danger that this trend may continue, especially given recent budget cuts in the public health sector.

We assign a beta prior distribution to represent the future uncertainty in the proportion of new ART patients who initiate TPT within 6 months of starting ART. We choose a prior mean of 40%, similar to the average value estimated by Thembisa over the 2010-24 period. We set the standard deviation to 15%, which yields 2.5 and 97.5 percentiles of the prior distribution of 13% and 71% respectively. This range is consistent with the range of IPT coverage estimates obtained from other South African studies, as noted previously.

#### **3.2 Efficacy of 3HP in sterilizing infection**

In previous modelling of the effect of isoniazid preventive therapy (IPT), we assumed that IPT in PLHIV provided protection only during the limited time that IPT was being taken. This was consistent with data from randomized trials conducted among African PLHIV, which found protection against TB rapidly waned after the discontinuation of IPT [24-26]. We argued IPT was unlikely to sterilize latent infection (Gavin Churchyard, personal communication). However, trials of IPT in immunocompetent individuals generally show effectiveness lasting over longer periods, with protection being insensitive to the duration of IPT beyond 12 months [27].

Unlike IPT, 3HP (3 months of isoniazid and rifapentine) is likely to be at least partially effective in sterilizing infection; experiments in mice show that rifapentine is substantially more effective than isoniazid in achieving long-term protection [28]. The WHIP3TB trial in African PLHIV found that 3HP provided similar benefits whether once-off or repeated [29], and an earlier trial of 3HR (similar to 3HP, but using rifampicin in place of rifapentine) in African PLHIV also found the impact was sustained over longer durations [26]. Randomized trials comparing 3HP/3HR to 6-9 months of IPT have found little difference in TB incidence between the two regimens in the first year of follow-up, but a divergence in effectiveness at longer durations, with 3HP/3HR appearing more effective at longer durations [26, 30, 31]. To model this dynamic, we assume that some fraction $\theta$ of latent TB infections are sterilized by 3HP (i.e. they return to the susceptible/uninfected state).

For the purpose of estimating $\theta$, we consider a simplified model for deriving overall trial efficacy from different efficacy parameters. Suppose $\lambda_{1}$ and $\lambda_{2}$ represent the annual incidence of TB due to fast progression and reactivation respectively, in the absence of any TPT. Over a two-year follow-up period in PLHIV, we would expect the number of incident TB cases following 9 months of IPT to be

$$\left( \lambda_{1}+\lambda_{2} \right)\left( N_{1}\left( 1-0.52 \right)+N_{2} \right)$$

where $N_{i}$ is the number of person years of observation in the *i*^th^ follow-up year, and 0.52 is the assumed efficacy of IPT during the year in which it’s taken [32]. (In specifying the effectiveness of IPT in Thembisa, we do not attempt to distinguish efficacy in preventing recurrent and fast-progression TB, due to the lack of data to delineate these effects.) If we provided the same cohort with 3HP instead, then the expected number of incident TB cases over two years of follow-up would be

$$\lambda_{1}{\left( N_{1}\left( 1-\alpha\right)+N_{2} \right)+\lambda}_{2}\left( 1-\theta\right)\left( N_{1}+N_{2} \right)$$

where $\alpha$ is the average reduction in fast-progression TB due to 3HP in the first 12 months following the start of 3HP (and we assume there is no lasting protection against fast progression after 12 months). In contrast to the model for IPT, here we are attempting to distinguish effects for reactivation and fast progression. Suppose the numbers of person years of observation are roughly equal in the two years of follow-up; then the ratio of the number of TB cases in the 3HP cohort to the IPT cohort, over two years, would be

$$\frac{\lambda_{1}{\left( 2-\alpha\right)+\lambda}_{2}\left( 1-\theta\right)\times2}{\left( \lambda_{1}+\lambda_{2} \right)\times1.48}$$

We have used two years here as it is close to the average follow-up duration in an early Ugandan trial comparing 6 months of IPT to 3HR and 3HRZ (adding pyrazinamide to 3HR) [26]. In this trial the relative risk of TB in the pooled 3HR/3HRZ arms compared to the IPT arm was 0.67 (95% CI: 0.42-1.08). If we assume roughly half of incident TB in PLHIV is due to reactivation [33, 34] (i.e. $\lambda_{1}$ = $\lambda_{2}$), and set the above equation to 0.67 (the same as the relative risk in the Ugandan trial), then

$$0.67=\frac{2-0.5\alpha-\theta}{1.48}$$

If we set $\alpha$ = 0.52 (the same as assumed for IPT), then solving this equation yields $\theta$ = 0.75, and if we replaced 0.67 with the upper and lower limits of 95% confidence interval for the RR (1.08 and 0.42 respectively), the $\theta$ estimate changes to 0.14 and 1.12 respectively. If we instead assumed that TPT was less effective in preventing fast progression than in preventing reactivation (as is often assumed in practice), we would need to assume higher values of $\theta$ in order to match the observed trial outcome. The same would be true if we assumed a higher proportion of incident TB due to fast progression of recently acquired TB. One could argue that these estimates of $\theta$ are conservative because 3HP is likely to be more effective in sterilizing infection than 3HR [28].

In the PREVENT TB trial, which compared 9 months of IPT and 3HR, over three years, the relative rate of TB incidence in the 3HP arm to that in the IPT arm was 0.38 (95% CI: 0.15-0.99) [31]. This trial was conducted predominantly in the US and Canada, where rates of TB transmission are believed to be very low, and most TB incidence is therefore likely to be due to reactivation. It would also appear that IPT provides more lasting protection in immunocompetent individuals [27], which comprised the vast majority of trial participants in this trial. If we set $\lambda_{1}$ = 0 and assume IPT provides lasting protection, and substitute the observed relative risk into the above equation, we obtain

$$0.38=\frac{\lambda_{2}\left( 1-\theta\right)}{\lambda_{2}\left( 1-0.52 \right)}$$

which implies $\theta$ = 0.82. Substituting the upper and lower limits of the 95% CI for the relative risk into this equation, we get lower and upper bounds of 0.52 and 0.93 respectively for $\theta$. This is not dissimilar from the estimate obtained using the Ugandan trial data. Other randomized trials have found less clear benefit of 3HP relative to IPT [35-37], though this may be a reflection of high rates of $\lambda_{1}$ in other settings. It should also be noted that almost all studies comparing 3HP and IPT have involved directly observed 3HP rather than self-administered 3HP [36], and it is not clear if similar efficacy would be achieved when 3HP is self-administered. Recognizing the wide uncertainty around these estimates, we represent the uncertainty around the value of $\theta$ using a vague prior (i.e. uniform (0,1)). The prior mean of 0.5 is lower than the estimates of 0.75 and 0.82 derived from the two previously cited trials [26, 31] because other trials have been more ambiguous regarding the relative efficacy of 3HP [35-37].

#### **3.3 Uptake of 3HP/TPT in household contacts**

Historically, TPT eligibility was limited to people living with HIV and household contacts under the age of 5 years [38]. More recently, in 2023, TPT eligibility was expanded to other high-risk groups, including all TB contacts (not just child household contacts), pregnant women, prisoners, health workers, and people who previously had TB [39]. Based on the most recent programme data for 2023 and 2024, we estimated that TPT uptake in household contacts was only 5.8% in 2023-24 and 7.7% in 2024-25 [5].

Studies that have assessed levels of TPT in child contacts of TB patients have generally found very low levels of uptake in South Africa, around 12-27% (Table S1). Frequently cited reasons for low uptake include healthcare providers not being aware of the recommendation to provide TPT to household contacts, or not telling parents to bring their children to the clinic for evaluation, and adults not bringing their children for evaluation (often related to inter-household movements) [40, 41]. However, in prospective South African studies in which additional efforts were made to trace child contacts and promote TPT uptake, initiation rates of around 75% have been achieved [42-44], which suggests that higher rates of TPT are possible. It is also possible that with better monitoring systems, high rates of TPT uptake may be achieved [45]

Table S1: Proportion of child household contacts (aged <5 years) who initiated TPT, in evaluations of routine care in South Africa

| Study | Location | # child  contacts^*^ | # starting  TPT | % starting  TPT |
| --- | --- | --- | --- | --- |
| Van Wyk *et al* [46] | Cape Town | 25^†^ | 3 | 12% |
| Van Wyk *et al* [47] | Cape Town | 24 | 5 | 21% |
| Philip and Feucht [40] | Tshwane | 28 | 6 | 21% |
| Thind *et al* [44] | Rustenburg | 552 | 125 | 23% |
| Osman *et al* [48] | Cape Town | 525 | 141 | 27% |

* Excluding children who were not eligible (either because they had TB or symptoms suggestive of TB, or because they were aged 5 years or older). † Including only children with folders that could be located (the study reports a lower % starting TPT because it included in the denominator children whose folders couldn’t be located).

We base our assumptions about future TPT uptake in adult household contacts on these studies. The uncertainty in the future proportion of adult household contacts who receive TPT ($P\left( t \right)$ for *t* ≥ 2027) is represented using a beta prior distribution with a mean of 0.25 and a standard deviation of 0.2. The mean is chosen to correspond roughly to the historic estimates of TPT uptake in South African child contacts of TB patients (Table S1). The 97.5 percentile of the distribution is 0.72, roughly consistent with the 75% that we consider to be a ‘best case’ scenario, based on the previously cited studies with enhanced TPT promotion [42-44]. The 2.5 percentile of the distribution is close to zero, and thus corresponds to a scenario in which there is minimal implementation of the new TPT recommendation.

Suppose $T\left( t-1 \right)$ is the number of adults starting TB treatment in month $t-1$, $h$ is the average number of adult household contacts who can be reached per newly treated TB patient, $\pi$ is the TB prevalence in household contacts, and $U\left( t \right)$ is the number of adults with untreated TB at the start of month $t$. We assume that there is an average delay of 1 month between an index case starting treatment and their household contact starting TPT. Then the probability that an individual who does not have TB in month $t$ starts TPT as a result of being a household contact is approximated as

$$\frac{T\left( t-1 \right)h\left( 1-\pi\right)P\left( t \right)}{N\left( t \right)-U\left( t \right)}$$

where $N\left( t \right)$ is the total population aged 15 and older. (Strictly speaking, the number currently on TB treatment should also be deducted from the denominator, but since the number treated is small relative to the total population size, it makes little difference to the approximation.) Assumptions about $h$ and $\pi$ are described in section 4.2. $P\left( t \right)$ is assumed to increase linearly from 0.077 in 2024 to the ultimate value in 2029 (the value sampled from the beta prior) and remain constant thereafter.

#### **3.4 Nutritional support**

Our model assumes that there is an increased TB risk associated with being underweight. However, this may be an over-simplification, as some studies suggest a more continuous relationship between BMI and TB risk [49, 50]. In the RATIONS trial, household contacts of TB patients had a 39% reduced risk of TB if they received nutritional support compared to controls [51]. Interestingly, the *proportional* reduction in TB risk was greater among those who had normal/high BMIs at baseline than in those who were underweight, although the absolute reduction in TB risk was similar in the two groups. This is consistent with the notion that gains in nutrition are almost always positive (in terms of reducing TB risk), regardless of the baseline weight.

Rather than model the effect of the RATIONS intervention as a reduction in the prevalence of under-nutrition, we assume a proportional reduction in TB incidence among all individuals who receive the nutrition support. To represent the uncertainty regarding the proportional reduction in TB incidence, we assign a beta prior with a mean of 0.39 and a standard deviation of 0.13. This distribution has 2.5 and 97.5 percentiles of 0.16 and 0.66 respectively, similar to the 95% CI reported for the effectiveness of the RATIONS intervention (0.15-0.57 [51]). We further allow for uncertainty regarding the likely timing of the introduction of nutritional intervention. The year of introduction (in years after 2025) is modelled using a Weibull distribution with a median of 10 years and a shape parameter of 0.55. As noted in the main text, this implies a 30% chance of the policy change occurring within 3 years and a 50% chance of the policy change occurring within 10 years.

We assume that the nutritional support is limited to household contacts of people with TB. We make the same assumptions about numbers of household contacts as in the modelling of household contact screening (see section 4.2). The nutritional support is assumed to be provided for 24 months, roughly in line with the average duration of follow-up in the RATIONS trial [51]. It is assumed that nutrition support reaches 80% of all eligible household contacts.

### **4. TB testing and diagnosis**

#### **4.1 Percentage of people seeking care for TB symptoms who are microbiologically tested**

As described in section 6.1.6 of the Thembisa TB report [5], we specify an ‘ultimate’ fraction of patients seeking treatment for TB symptoms who get microbiologically tested, which applies for all years from 2028. South African studies have estimated this proportion to be low: between 3% and 84% (with a median of 30%) in studies conducted over the 2011-2016 period [52-55]. However, the Thembisa model estimates that the rate of testing has declined in recent years, as testing volumes have dropped; over the period from mid-2016 to mid-2024, the model estimates only 12.4% of individuals seeking treatment for TB symptoms were microbiologically tested [5].

We represent the uncertainty in this parameter using a beta distribution with a mean of 0.125 and a standard deviation of 0.065. The mean is in line with the Thembisa average over the 2016-24 period (i.e. assuming no major future changes to testing in symptomatic individuals), while the standard deviation was chosen such that the 2.5-97.5 percentile range of the prior distribution (0.03-0.28) corresponds with the lower limit and median estimates in South African studies conducted when testing volumes were substantially higher [52-55]. For the years 2024-2027, the proportions of TB patients tested are linearly interpolated between the proportion estimated for 2023-24 (based on programme data) and the sampled value for 2028 and subsequent years.

One possible solution to the problem of low rates of testing in symptomatic individuals is to introduce tongue swab testing to replace sputum testing in symptomatic individuals. This would have several advantages. Firstly, tongue swab specimens are easier to collect than sputum specimens; many patients are either unable to produce sputum or produce only saliva [56], and both patients and providers report that tongue swabs are more acceptable [57]. Secondly, recently developed tests are near-point-of-care and can be performed in primary health care facilities with minimal equipment and provider training [58]; the tests can be performed on tongue swabs or sputum. Thirdly, the tongue swab tests are likely to be substantially cheaper than the existing sputum-based tests [59]. The sensitivity and yield of tongue swab testing appears to be similar to that of sputum testing in symptomatic individuals [58, 60], although sensitivity may be relatively poor in asymptomatic individuals (see section 4.7). There is thus likely to be a substantial improvement in the microbiological testing rate, as a result of overcoming healthcare workers’ concerns about the cost and time required to perform sputum-based tests in reference laboratories [61]. With point-of-care HIV testing, for example, it has been possible to achieve testing coverage levels of over 90% in pregnant women [62] and TB patients [63-65], high priority groups in which South African guidelines have recommended regular testing [66]. We therefore assume that if tongue swab testing were introduced and recommended in national guidelines, the proportion microbiologically tested could increase to as much as 90%. We represent the uncertainty in this proportion using a beta prior distribution with mean of 0.51 and standard deviation of 0.21; these parameters were chosen such that the 2.5 percentile (0.12) would correspond to the current ‘baseline’ estimate of the coverage of microbiological testing (i.e. conservatively assuming that the introduction of tongue swab testing would not lead to any improvement in levels of microbiological testing), and the 97.5 percentile (0.89) would roughly correspond to the likely upper bound of 90% coverage (the mean of 0.51 is the average of the upper and lower bounds).

To represent the uncertainty around the timing of the potential introduction of tongue swab testing, we assign a Weibull prior distribution to the time (in years after 2025), with a median of 10 years and a shape parameter of 0.55 (the same distribution as for other potential new interventions).

#### **4.2** **Active case finding/TUTT uptake in contacts of TB patients**

The probability that an individual with untreated TB at the start of month $t$ is screened for TB as a result of active case finding or TUTT is approximated as

$$d_{1}\left( t \right)=\frac{T\left( t-1 \right)hS\left( t \right)\pi}{U\left( t \right)}$$

where $T\left( t-1 \right)$ is the number of adults starting TB treatment in month $t-1$ (having been diagnosed through passive case finding), $h$ is the average number of household contacts who can be reached per newly treated TB patient, $S\left( t \right)$ is the probability of a household contact being screened in month $t$, $\pi$ is the TB prevalence in household contacts, and $U\left( t \right)$ is the number of adults with untreated TB at the start of month $t$. The implicit assumption made here is that there is an average delay of 1 month between an index case starting treatment and their household contact being screened (and starting treatment). Strictly speaking, $\pi$ should be defined as the prevalence of *untreated* TB in household contacts, but since the proportion of prevalent TB that is treated is low [67, 68], it makes little difference to the approximation.

Similarly, the probability that an individual who does not have TB in month $t$ is screened for TB as a result of active case finding or TUTT is approximated as

$$d_{0}\left( t \right)=\frac{T\left( t-1 \right)hS\left( t \right)\left( 1-\pi\right)}{N\left( t \right)-U\left( t \right)}$$

where $N\left( t \right)$ is the total population aged 15 and older. (Strictly speaking, the number of treated individuals should be deducted from the denominator, but since the number treated is small relative to the total population, it makes little difference to the approximation.) It is worth noting that generally we would expect $d_{1}\left( t \right)>d_{0}\left( t \right)$, since people who have TB are more likely to have household contacts with TB than people without TB.

We set the assumed average number of household contacts screened ($h$) to 3.0, based primarily on three South African studies that found means of 2.9 [69], 3.0 [67] and 3.2 [70]. This may overstate the parameter we are estimating because our definition implicitly excludes children. However, other South African studies have found higher average numbers of household contacts (e.g. 5.0 [44] and 6.2 [71]). It is also worth noting that many household contacts (especially children) are unable to produce sputum, and so estimates of average numbers of sputum specimens from household contacts are typically lower, e.g. 1.8 [70], 2.0 [72] and 0.3 [73].

A number of South African contact studies have estimated the TB prevalence in household contacts of index TB patients ($\pi$). However, it is important to take into account that the methods used in these studies typically have poor sensitivity, and thus under-estimate the true TB prevalence. Table S2 shows the reported and adjusted TB prevalence estimates from these studies, where the latter are based on assumed levels of test/algorithm sensitivity. The pooled prevalence estimate, using a random effects model, is 7.0% (95% CI: 3.9-10.0%). This may overstate the true prevalence because the denominator is contacts who could provide a sputum specimen, but those unable to produce sputum are less likely to have TB. A further limitation of our approach is that it implicitly assumes the prevalence of TB in household contacts is constant over time. Although this may be untrue, a crude analysis of the data in Table S2 does not suggest any trend in TB prevalence over time. We do not have enough data to suggest a better approach to incorporating time-varying TB prevalence, but it might be expected that if TB transmission rates decline in future, $\pi$ will also decline.

Table S2: Proportion of South African TB household contacts who have TB

| Study | Period | Diagnosis | Assumed | n | TB prevalence | |
| --- | --- | --- | --- | --- | --- | --- |
|  |  |  | sensitivity |  | Reported | Adjusted |
| Velen *et al* [69] | 2013-2015 | Culture if WHO 4SS+^a^ | 71%^a^ | 356 | 5.9% | 8.3% |
| Thind *et al* [44] | 2009-2011 | Culture if symptomatic^b^ | 35.1%^b^ | 3029 | 3.1% | 8.7% |
| Shapiro *et al* [67] | 2009 | Culture | 100% | 2147 | 7.8% | 7.8% |
| Lebina *et al* [70] | 2011-2012 | Culture/Xpert^c^ | 100/65%^d^ | 1378 | 2.2% | 2.6% |
| Deery *et al* [73] | 2012 | Xpert only | 65%^d^ | 25 | 4.0% | 6.2% |
| Gilpin & Hammond [72] | 1984 | Microscopy only | 35.5%^e^ | 132 | 3.0% | 8.5% |
| Pooled |  |  |  |  |  | 7.0% |

^a^ WHO 4SS+ = WHO 4-symptom screen positive [74]. ^b^ Corresponds to chronic cough symptoms [75]. ^c^ Different trial arms received culture or Xpert testing. ^d^ Assuming Xpert testing is 65% sensitive, which is the average sensitivity of Xpert/Ultra in the context of active case finding [76]. ^e^ Weighted average proportion of South African TB cases that are smear-positive in household-based studies.

The last parameter that we consider is the proportion of household contacts who are screened, $S\left( t \right)$. This takes into account the difficulty of taking sputum specimens: the proportion of household contacts who are able to produce sputum specimens has been estimated at 56% [70] and 39% [77] in South African studies. This suggests an upper bound on the $S\left( t \right)$ parameter, although it is possible that individuals with TB may be more likely to produce sputum. As noted in section 3.3, the new TPT policy in South Africa is to provide TPT to all household contacts of TB patients [39], and this implies a policy of routinely screening household contacts for TB (which we estimated to have reached 7.7% of household contacts in 2024-25 [5]). We define $k$ as the future proportion of household contacts who receive screening (at a minimum, symptom screening followed by sputum testing in those who report symptoms). We use a uniform (0, 0.6) prior to represent the uncertainty in the $k$ parameter, based on the previously noted difficulties in collecting sputum. The scaling up of household contact screening is assumed to occur linearly, starting from zero in 2024-25 to the maximum value by 2029-30.

The probability that a screened individual with TB is diagnosed depends on what tests are conducted and what symptoms they have. For a household contact who has TB, with smear status *m*, the assumed sensitivity of the diagnostic algorithm is $\eta_{m}$. This is calculated as

$$\eta_{m}=U_{m}\left( p_{m}+\left( 1-p_{m} \right)J \right)$$

where $U_{m}$ is the sensitivity of the Xpert Ultra test, $p_{0}$ and $p_{1}$ are the proportions of smear-negative and smear-positive TB patients respectively who have symptoms, and $J$ is the proportion of screened individuals who receive ‘targeted universal testing for tuberculosis’ (TUTT, i.e. Xpert testing is conducted regardless of symptoms [78]). The $p_{0}$ and $p_{1}$ parameters are fixed at the default Thembisa values (0.20 and 0.60 respectively). We represent the uncertainty in the $J$ parameter using a uniform (0, 1) prior distribution. The $U_{0}$ and $U_{1}$ parameters are set to 0.77 and 0.99 respectively, the same as assumed in Thembisa for passive case finding [79]. Although it could be argued that the sensitivity of Xpert Ultra is likely to be lower in the context of active case finding, reviews of Xpert Ultra sensitivity in the context of active case finding do not report estimates disaggregated by smear status [76]. Since our model is already accounting for the higher smear-negative proportion in active case finding compared to passive case finding, and this is likely to explain much (if not all) of the lower sensitivity, we do not consider it necessary to make different assumptions about the sensitivity of Xpert Ultra between active and passive case finding.

#### **4.3 TUTT uptake in PLHIV**

Historically, TB policy in South Africa was to test people living with HIV (PLHIV) only if they had TB symptoms. However, the new policy, targeted universal testing for TB (TUTT), requires testing all PLHIV for TB, regardless of their symptoms [78]. Although this may be effective in diagnosing TB earlier, it is debatable whether it would be feasible to do this at every PLHIV healthcare visit. It is also debatable whether it would be cost-effective, particularly given that there are already (even in the absence of TUTT) extremely high rates of TB testing in South African PLHIV [80]. Indeed, in the original TUTT trial, PLHIV were the risk group with the lowest TB positivity rate [81], which suggests that it would be more efficient to target TUTT to other risk groups. TUTT has been declared national policy,^[1]^ although it is not clear to what extent it is being implemented, as testing statistics are not disaggregated according to testing modality/reason for testing.

In our model, we assume for simplicity that TUTT for PLHIV is provided only to those who are on ART (although it is theoretically possible that PLHIV who are not on ART might be offered TUTT, it is unlikely that someone would be in HIV care if they are not on ART). For an individual with TB (of smear status *s*) who is on ART, the monthly rate of starting TB treatment due to TUTT in ART patients is

$$\frac{f_{H}\left( t \right)}{12}\left[ T\left( t \right)+\left( 1-T\left( t \right) \right)c_{s} \right]{Se}_{s}\left( 1-L \right)$$

where $f_{H}\left( t \right)$ is the average annual number of times an ART patient is screened for TB in year *t*, $T\left( t \right)$ is the proportion of actively screened individuals who are screened according to TUTT protocols (the remainder are tested only if they are symptomatic, with $c_{s}$ representing the prevalence of symptoms), ${Se}_{s}$ is the sensitivity of the Xpert test, and *L* is the initial loss to follow-up probability in individuals who are diagnosed positive. (The symptom prevalence, sensitivity and initial loss to follow-up assumptions are described more fully in sections 6.1.2, 6.1.4 and 6.2 respectively of the Thembisa TB report [5].) For the sake of simplicity, we assume here that $T\left( t \right)$ = 1 in all years in which $f_{H}\left( t \right)>0$. In initial discussions with the TB Think Tank and National Department of Health, to assess what might be achieved under the current National Strategic Plan [1], it has been suggested that a target of around 3 million PLHIV tested per annum would be reasonable [82]; this is equivalent to roughly half of all ART patients in South Africa. In a best case scenario, it might be hoped that all ART patients would be actively screened once per annum, but realistically even the target of 3 million tests per annum is extremely ambitious (since this would more than double the total number of TB tests performed in South Africa each year). In our attempt to calibrate the Thembisa TB model to the most recent programme data, we estimated 0.22 tests per year in ART patients [5], but there is a high degree of uncertainty around this because of the previously noted lack of disaggregation in national testing statistics. We represent the uncertainty in $f_{H}\left( t \right)$ for years 2030 and later using a gamma distribution with a mean of 0.30 and a standard deviation of 0.25. The mean of 0.3 is slightly more conservative than the target of roughly half of all ART patients, but more optimistic than our current baseline estimate of 0.22. The 2.5 and 97.5 percentiles are 0.02 and 0.95 respectively, wide enough to reflect substantial uncertainty. The 97.5 percentile corresponds roughly to what might be accomplished in a best case scenario, where every ART patient is actively screened for TB once per annum.

#### **4.4 TUTT in previously treated TB patients**

As noted in section 4.3, TUTT has been declared national policy in South Africa, and this includes actively screening for TB in patients who have had TB in the last two years. The population who have recently been treated for TB is substantially smaller than the population of people on ART, and correspondingly the targeted numbers of TB tests in recovered TB patients are lower, at around 300 000 per annum [82].

In our model, we assume for simplicity that TUTT for previously treated individuals diagnoses only those TB cases that develop in the ‘high-risk’ phase following TB recovery. This high-risk phase is assumed to last for 6 months on average, and people are assumed to experience a high risk of relapse during this phase (as described in section 6.3.4 of the Thembisa TB report [83]). Because the ‘low-risk’ phase is associated with a low risk of relapse, and because people who develop TB in the low-risk phase are less likely to get diagnosed within the 24-month window, our model simulations suggest they account for only about 20% of the TB cases that would be diagnosed within 24 months after recovery – suggesting it is probably reasonable to ignore the low-risk phase for the purpose of modelling TUTT in previously treated individuals.

Similar to the approach described for TUTT in people living with HIV, we assume that for an individual with TB (of smear status *s*), the monthly rate of starting TB treatment due to active case finding in previously treated patients is

$$\frac{Z\left( t-1 \right)f_{P}\left( t \right)}{12}\left[ T\left( t \right)+\left( 1-T\left( t \right) \right)c_{s} \right]{Se}_{s}\left( 1-L \right)$$

where $Z\left( t \right)$ is the proportion of TB cases in year *t* that occur in people in the high-risk post-TB phase, $f_{P}\left( t \right)$ is the average annual number of times a previously treated patient is screened for TB in year *t*, and the remaining terms are the same as defined in section 4.3 ($T\left( t \right)$ is the proportion of actively screened individuals who are screened according to TUTT protocols, ${Se}_{s}$ is the sensitivity of the Xpert test, and *L* is the initial loss to follow-up probability in individuals who are diagnosed positive). The $Z\left( t-1 \right)$ term is calculated from the model outputs in the previous year. The $f_{P}\left( t \right)$ term is estimated by noting that our current projection of the average annual numbers of adults treated for TB, over the 2025-2030 period is about 238 000 per annum, and our projection of the TB deaths on treatment, over the same period, is around 22 000 per annum, implying around 216 000 adults surviving to the end of TB treatment per annum. If around 300 000 people are screened in the 24 months after completing treatment, each year (in line with current targets), that would imply a $f_{P}\left( t \right)$ value of 0.69 (300 000/(2 × 216 000), where the 2 is accounting for the 2 years that people remain eligible after recovery). Although the 300 000 target is perhaps more achievable than the 3 million target for people on ART, in terms of TB testing capacity, people with prior TB are more difficult to reach (because they are not an easily identifiable group in clinical settings). We therefore represent the uncertainty in $f_{P}\left( t \right)$ for years 2029 and later using a gamma distribution with a mean of 0.50 and a standard deviation of 0.25. The mean is slightly lower than the implied target of 0.69, to reflect the challenges associated with identifying recently treated TB patients, but we have chosen the standard deviation of 0.25 to give a wide uncertainty range (2.5 and 97.5 percentiles are at 0.14 and 1.10 respectively).

#### **4.5 Rollout of door-to-door TB screening (symptoms)**

Three recent trials have assessed the impact of door-to-door TB screening in resource-limited settings: one in Zimbabwe [84], one in Vietnam [85] and one in Malawi [86]. The first two involved regularly offering TB screening – at three annual intervals in Vietnam and at six 6-monthly intervals in Zimbabwe – while the Malawian trial involved a once-off TB screen. In the Vietnam trial, the proportion of adults who consented to participate and who were capable of providing sputum samples declined from 45% in the first round to 35% in the third round. (Since only approximately half of asymptomatic individuals are able to produce sputum [70, 77], the proportion actually reached by the screening teams may have been close to 100% in round 1.) In the Zimbabwe trial, the cumulative proportion of the adult population reached was only 8.2% (rates were not reported separately for each interval), although the proportion tested was found to be higher in communities with higher testing yields (suggesting that uptake is proportionally higher when targeted to high-prevalence communities) [84]. In the Malawian trial, the proportion of the population reached was not directly reported, although the authors estimated that only 23% of people with cough symptoms submitted sputum, which would suggest a participation rate of 30% if the sputum production rate in symptomatic individuals is 77% [56].

We define $\delta$ as the annual rate at which individuals are reached by household screening teams and consent to screening. We specify a gamma prior distribution to represent uncertainty around $\delta$ (assuming this intervention is introduced). The prior distribution mean is set to 0.30 (the median estimate from the three previously cited trials) and the standard deviation is set to 0.23. With these parameters the 2.5 percentile of the prior distribution is 0.028 per annum (roughly consistent with the annualized rate derived from the Zimbabwean trial [84]), and the 97.5 percentile is 0.89, roughly consistent with the annual rate of nearly 100% observed in the first round of the Vietnam trial [85].

For an individual with smear-positive TB, the monthly rate at which they are diagnosed through door-to-door screening, if sputum-based testing is conducted only in people who are symptomatic, is calculated as

$$\frac{\delta}{12}c_{1}U_{1}Se_{1}$$

where $c_{1}$ is the proportion of smear-positive individuals who are symptomatic (i.e. they would automatically get Xpert testing), is the proportion of symptomatic individuals who are able to produce sputum, and $Se_{1}$ is the sensitivity of the Xpert test in smear-positive individuals. Similarly in people who have smear-*negative* TB, the monthly rate of diagnosis through door-to-door screening is

$$\frac{\delta}{12}c_{0}U_{1}Se_{0}$$

where $c_{0}$ and $Se_{0}$ are the symptomatic proportion and sensitivity of Xpert in smear-negative TB respectively. The assumptions about symptomatic proportions and Xpert sensitivity are specified in sections 6.1.2 and 6.1.4 of the Thembisa TB report [5]. In contrast to the low rates of sputum production noted previously in asymptomatic adults, high rates of sputum production have been estimated in South African studies of mostly symptomatic individuals (61% [87] and 86% [88]); we therefore set $U_{1}$ to 0.74, the median of these estimates (and similar to the results of an international meta-analysis, which found the proportion to be 77% [56]).

Because door-to-door screening is not current policy in South Africa, we specify a prior distribution to represent the uncertainty around the time to introducing this policy in South Africa. As with other new policy timing parameters, the year of introduction (in years after 2025) is modelled using a Weibull distribution with a median of 10 years and a shape parameter of 0.55. As noted in the main text, this implies a 30% chance of the policy change occurring within 3 years and a 50% chance of the policy change occurring within 10 years. A major constraint on the rollout of door-to-door testing is likely to be the substantial cost of doing so; if the intervention is introduced, it is likely to be targeted to communities in which there is a high prevalence of untreated TB (e.g. urban informal settlements), and indeed the WHO only recommends community-based screening when the prevalence of untreated TB exceeds 0.5% [74].

#### **4.6 Rollout of door-to-door TB screening with digital chest X-ray (dCXR)**

The 2023-2028 National Strategic Plan specifically mentions digital chest X-ray (dCXR) as one of the tools that should be used to improve levels of screening [1]. Combined with computer-aided detection (CAD), dCXR technology could potentially reduce human resource requirements (specialist time required to read and interpret chest X-rays). However, there are concerns regarding the scalability of dCXR screening, given that the portable units are expensive and need to be operated by skilled radiographers.

The Vukuzazi study in rural KwaZulu-Natal did not involve door-to-door testing, but did involve mobile vans from which TB screening could be accessed. Over two years, 50% of the adult population enrolled for testing [89]. In the first year specifically, 9 914 out of 36 097 adults received chest X-ray screening (27%) [90], which is similar to the assumed mean annual rate of door-to-door screening in the previous section (0.30).

Given the uncertainty around the feasibility of introducing dCXR at scale, we assign a vague prior (uniform on the interval [0, 1]) to represent the uncertainty in the proportion of door-to-door screens in which dCXR screening is offered to individuals who do not have TB symptoms. We assume the same sensitivity and specificity of dCXR as estimated in a recent South African study in rural KwaZulu-Natal (83% and 63% respectively [90]). This means that for an individual with smear-positive TB, the monthly rate at which they are diagnosed, through door-to-door screening, is

$$\frac{\delta}{12}\left( c_{1}U_{1}Se_{1}+\left( 1-c_{1} \right)D\times0.83{\times U}_{0}Se_{1} \right)$$

where $D$ is the proportion of households in which digital chest X-ray is offered to people who are asymptomatic, and $U_{0}$ is the proportion of asymptomatic individuals who are able to provide sputum. Similarly in people who have smear-negative TB, the monthly rate of diagnosis through door-to-door screening is

$$\frac{\delta}{12}\left( c_{0}U_{1}Se_{0}+\left( 1-c_{0} \right)D\times0.83\times U_{0}Se_{0} \right)$$

Our implicit assumption is thus that the sensitivity of digital chest X-ray is the same in smear-negative and smear-positive TB. As noted previously, we represent the uncertainty in $D$ using a uniform (0, 1) prior distribution. South African studies in predominantly asymptomatic individuals have estimated that only 39% [77] and 57% [70] of individuals are able to produce sputum; we therefore set $U_{0}$ to 0.47, the median of these two values.

We have focused here only on the use of dCXR in community-based screening. Although there have also been proposals to introduce dCXR screening in health facilities, it appears this would yield relatively few additional diagnoses, relative to existing symptom screening [87] or relative to LAM testing in PLHIV [91]. Although the question of dCXR use in public health facilities has been discussed by the South African TB Think Tank, there has not yet been any consensus on how best to deploy this technology, if at all (Limakatso Lebina, personal communication). Since it is unclear how to define appropriate ‘high risk’ groups for targeted screening, when most of these high-risk group are already covered by TUTT, we have not attempted to model this scenario of dCXR use in health facilities.

#### **4.7 Tongue swabs in the context of door-to-door testing**

As noted previously, a major challenge in active case finding and community-based testing is that many individuals struggle to produce sputum, especially those who are asymptomatic. Molecular tests can also be performed on tongue swabs, which although less sensitive than on sputum [92], are preferable to not offering any test. Most individuals prefer providing tongue swab specimens to providing sputum [57], and with the recent introduction of the MiniDock device, there is the added advantage that the test can be performed on a near-point-of-care basis with minimal equipment [58]. Previous studies in the context of HIV testing have shown that the switch from offering laboratory testing to offering point-of-care testing, and to offering simpler sample collection methods (e.g. from venous blood to saliva) can roughly double rates of testing uptake in community settings [93, 94]. The MiniDock device is also substantially cheaper than existing laboratory-based testing [59], making it likely that it would be more widely rolled out.

We therefore consider the potential introduction of tongue swab testing in the context of door-to-door testing, modifying the equations specified in the previous section. We define $ϒ$ as the proportion of communities reached by door-to-door testing in which tongue swab testing is offered. For the sake of simplicity, we assume that in these communities all individuals offered testing choose tongue swab testing over sputum testing. (In reality, some may prefer to provide sputum, and so we may under-estimate the impact of the intervention, although it is unlikely to be a major source of bias, given the higher acceptability of tongue swab testing [57].) Based on the evidence cited in the previous paragraph, we assume that the annual rate of uptake of TB screening is doubled (to $2\delta$) in these communities in which tongue swab testing is offered. Then the modified monthly rate at which a person with TB (with smear status *m*) is diagnosed through door-to-door screening is

$$\frac{ϒ2\delta}{12}\left( c_{m}Se_{m}R_{1}+\left( 1-c_{m} \right)Se_{m}R_{0} \right)+\left( 1-ϒ \right)\frac{\delta}{12}\left( c_{m}U_{1}Se_{m}+\left( 1-c_{m} \right)D\times0.83U_{0}Se_{m} \right)$$

$$=\frac{\delta Se_{m}}{12}\left\{ 2ϒ\left( c_{m}R_{1}+\left( 1-c_{m} \right)R_{0} \right)+\left( 1-ϒ \right)\left( c_{m}U_{1}+\left( 1-c_{m} \right)D\times0.83U_{0} \right) \right\}$$

where $R_{s}$ is the relative sensitivity of tongue swabs (when compared against Xpert on sputum) in people of symptom status *s*. The relative sensitivity of tongue swabs is difficult to estimate. In a recent systematic review of tongue swab sensitivity estimates, only one study estimated sensitivity in predominantly asymptomatic individuals (52%), and in the remaining seven studies that assessed sensitivity in mostly symptomatic individuals, the median sensitivity was 83% [92]. A recent analysis found an even higher relative sensitivity of 0.91 when using the MiniDock tongue swab test [58]. Based on this evidence we set $R_{0}$ to 0.52 and $R_{1}$ to 0.90.

It is not clear if tongue swab testing will be rolled out; it is possible that tests might be offered only in certain settings. We therefore represent the uncertainty in the $ϒ$ parameter using a uniform (0, 1) prior distribution.

#### **4.8 Culture testing if negative on initial TB test**

Although culture is generally considered the gold standard in TB diagnosis, culture tests take a long time to deliver results. For this reason, they tend to be used only in special circumstances – current guidelines recommend use of culture only if the initial Xpert test result (on sputum) is negative and the patient is HIV-positive (historically culture testing was also recommended if the patient had previously been treated for TB). Even when these conditions are met, South African studies suggest relatively low levels of culture use [95, 96]. Our current default assumption is that culture testing is conducted in Xpert-negative people with TB symptoms only 30% of the time if they are HIV-positive. However, there is substantial uncertainty around these assumptions. Data from Cape Town suggest that use of culture testing in eligible individuals was relatively high (83%) in the period before Xpert was introduced, but dropped to 41% after the transition to Xpert testing [96]. On the other hand, it was found in the XTEND trial that only 11% of HIV-positive individuals who tested negative on Xpert were subsequently tested by culture [95] – this is 0.36 times our assumed default. We specify the parameter Z to represent the ratio of the future culture testing frequency (in 2028 and subsequent years) to that assumed in the period up to 2025. To represent the uncertainty in this parameter we assign a gamma prior with a mean of 1 (i.e. assuming no change from the default, on average) and standard deviation of 0.42. This distribution has 2.5 and 97.5 percentiles of 0.36 and 1.98 respectively, the former being consistent with the conservative multiple estimated from the XTEND trial data, and the latter being more compatible with the higher testing rates in the Cape Town study.

#### **4.9 Relative rate of TB screening in patients seeking treatment for TB symptoms**

In theory, all patients who are attending health facilities and experiencing TB symptoms should receive TB testing, regardless of whether the symptoms are the reason for their healthcare visit. However, in many public health facilities, staff shortages and time pressures mean that patients do not get screened for other conditions. South African studies reporting on microbiological testing in individuals attending clinics with TB symptoms have estimated the probability of microbiological testing is between 2.2 and 10.8 times higher in patients who come to clinics because of those symptoms than in symptomatic patients attending health facilities for other reasons [52-54]. In reviewing these studies and in setting the default Thembisa assumptions, we noted that these multiples are highest at low levels of testing in patients seeking treatment for TB symptoms [83], implying a lower fidelity to testing guidelines when resources for testing are limited. We previously set the default value for the multiple, over the period after 2021, to 11.1, based on the relatively low levels of microbiological testing estimated in recent years.

In setting a prior distribution to represent uncertainty in future values of this multiple, we choose a gamma distribution with a mean of 11 (consistent with the current default) and a standard deviation of 6.5. This distribution has 2.5 and 97.5 percentiles of 2.2 and 26.9 respectively, the latter corresponding to the lower bound on the ratio measured in South African studies [53]. Although the upper bound is substantially higher than that measured in any South African studies, the few published studies were all conducted when rates of TB testing were relatively high, and given the substantial decline in TB testing rates over the last decade, higher multiples are quite possible for recent years (and into the future). Because of the negative correlation between the multiple and the rate of microbiological testing in people seeking treatment for TB symptoms (the parameter described in section 4.1), we assume a negative correlation coefficient (-0.5) between these two parameters.

### **5. TB treatment**

#### **5.1 Initial loss to follow-up**

One of the stated objectives of the 2023-2028 National Strategic Plan is to improve linkage to treatment after diagnosis, for example through counselling after TB diagnosis [1], or through better tracing of patients through electronic medical record systems. The NSP recognizes that linkage is particularly poor when diagnosis occurs in hospital settings. In the LINKEDin study, conducted across three South African provinces, a combination of improved hospital recording and patient tracing systems was found to reduce initial loss to follow-up (ILFTU), although impacts were heterogeneous across provinces – ranging from no impact in Gauteng to a 42% reduction in KwaZulu-Natal [97]. A South African pilot study also found that use of an mHealth application reduced ILTFU from 31% to 15%, although this reduction was not statistically significant due to small sample sizes [98]. Similarly, an SMS reminder intervention was found to slightly reduce ILTFU in South African patients diagnosed with TB [99]. The use of point-of-care diagnostics (currently under evaluation) could potentially further reduce ILTFU by removing the need for patients to return to a health facility to receive their TB diagnosis.

In the Thembisa TB model, the assumed current ILTFU probability is 17.6%, based on a meta-regression of South African studies that have estimated rates of ILTFU in TB patients [83]. However, there is substantial heterogeneity across individual studies, with the ILTFU rate in primary healthcare settings reaching as high as 29% in one study, conducted across five provinces [100, 101]. To represent the uncertainty regarding future ILTFU rates in South Africa, we use a beta prior distribution with a mean of 17.6% and standard deviation of 5%. This distribution has 2.5 and 97.5 percentiles of 9.0% and 28.4% respectively. The mean is thus the same as the current default assumption, while the 2.5 percentile corresponds to what might be expected in a best case scenario if a nationally-implemented intervention (such as the ones described in the previous paragraph) is 50% effective in reducing ILTFU. The 97.5 percentile corresponds to a worst case scenario in which ILTFU rates nationally are similar to those in the previously cited study in five provinces [100, 101].

The default assumptions about ILTFU in Thembisa relate to diagnosis in heathcare settings. However, there is concern that linkage to treatment is likely to be poorer when diagnosis occurs in community settings. The same phenomenon is observed in the case of HIV; for example, in a previous modelling study we assumed the rate of linkage to ART after an HIV diagnosis in community settings was only 0.68 times that when diagnosed in healthcare settings [102], based on ratios of 0.61-0.73 in African studies [103-106]. Although we lack similar comparisons for TB, local studies do confirm that rates of linkage are low in the context of household contact screening [67, 107]. We therefore represent the uncertainty around the relative rate of linkage for community-based testing using the same prior distribution as in our previous work, viz. a beta distribution with a mean of 0.68 and standard deviation of 0.15.

#### **5.2 Novel treatment regimens for drug-sensitive TB**

Standard therapy for drug-sensitive TB involves a six-month course of a combination of antibiotics. Because of the long duration of treatment, many patients drop out of treatment before completing treatment, reducing the chance of effective cure and increasing the risk of TB recurrence. In recent years there have been a number of trials to investigate shorter regimens for drug-sensitive TB. In one trial, four months of treatment with a drug combination that included rifapentine (rather than the standard rifampicin) was found to be non-inferior to the standard 6-month regimen, although TB-free survival at 12 months was marginally lower than in the standard of care trial arm [108]. In another trial, four months of rifampicin (at a higher dose than in the standard 6-month regimen) was found to be inferior to the standard 6-month regimen [109].

Based on the first of these trials, WHO has conditionally recommended the 4-month regimen [110]. However, many uncertainties remain regarding the recommendation. It is not clear if the recommendation is appropriate in TB patients with advanced HIV and more severe forms of TB, and there are concerns that there may be additional drug resistance testing required. There is also uncertainty regarding the cost and availability of rifapentine. Because of these uncertainties, South Africa has not yet changed its recommended regimen for drug-sensitive TB, although the 2023-28 TB strategic plan aims for the introduction of the shorter regimen [6], and the possibility of testing out the new regimen in a number of pilot sites is being considered.

Because it is not clear if and when South Africa will switch to the 4-month regimen, we represent the uncertainty in the time to introducing the new regimen using a Weibull distribution with a median of 10 years and a shape parameter of 0.55. The assumed relative effectiveness of the 4-month regimen is discussed in section 5.4.

#### **5.3 Treatment discontinuation rate**

The 2023-2028 National Strategic Plan outlines a number of strategies to improve treatment adherence and completion, including counselling services, treatment buddies, SMS reminders and systems to track and trace patients who have stopped taking treatment [1]. There is also some interest in using nutritional support and/or economic support to improve retention in care, given the economic barriers that many patients face in attending clinics regularly. Randomized trials of nutritional interventions have not produced clear evidence of better TB treatment completion [111, 112]. However, one cash transfer trial found a significant effect on treatment success (OR 1.6, 95% CI: 1.0-2.6) [113], and other observational studies in Latin America found that cash transfers were associated with significantly increased treatment success [114] and reduced treatment default (RR 0.36, 95% CI: 0.23-0.57) [115].

Although our model makes allowance for the implementation of DOTS (directly observed treatment, short-course) in the period before 2020, we have assumed only a modest impact of DOTS on treatment discontinuation rates – in part because of poor adoption of DOTS and in part because DOTS places a heavy burden on patients in the local setting [83]. However, it has been argued that alternative models could achieve similar or better outcomes. For example, in a systematic review and meta-analysis, DOTS delivered in community settings achieved better treatment completion than DOTS in clinic settings (OR 2.92, 95% CI: 1.15-7.41), though this was based on a single RCT and was not replicated in observational cohort studies [116].

Patient education interventions and support groups could also significantly improve completion rates. In the same systematic review, patient education significantly improved completion rates (OR 1.72, 95% CI: 1.32-2.22), but again based on a single RCT [116]. Two studies of patient support groups found increased treatment completion (RRs of 1.20-1.47) and reduced LTFU (RRs of 0.31-0.50) [116].

Although there is potential for new interventions to increase treatment completion and reduce rates of default, these need to be weighed against the potential for declining retention in care in future. A number of studies have found that patients are more likely to default TB treatment if they have fewer TB symptoms [117, 118], or if they experience a more rapid resolution of symptoms [119]. This means that as TB programmes transition towards greater screening in asymptomatic individuals, a decline in average TB treatment completion rates might be expected [6]. In one African trial, for example, TB symptom scores ranged between 1 and 12 (median 5), and the odds of missed visits decreased by a factor of 0.64 for each unit increase in the TB symptom score [117]. In another South African trial, the average number of symptoms in 147 TB patients who reported cough of >2 weeks duration (the primary symptom screen currently used in South Africa) was 3.6, compared to 1.6 in all 702 TB patients [120]. The results of these two trials imply that if treatment initiation was in predominantly asymptomatic TB cases, the odds of treatment default could be as much as 2.44 times (0.64^(1.6-3.6)^) the current odds, in a context where patients are initiated on treatment based primarily on reporting of prolonged cough.

As described in section 6.3.2 of the Thembisa TB model report [5], we have estimated rates of treatment discontinuation based on data from the Electronic TB Register (ETR) over the 2004-2016 period, but have allowed for a slight increase in these rates over the 2016-21 period due to DOTS no longer being implemented. The base annual rates of treatment discontinuation, in the period after 2021, are 0.418 for males and 0.320 for females. It is not clear to what extent these ‘base rates’ may change in future years, as a result of new interventions. We specify a multiplier to these base rates to obtain the rates that apply from 2029 onwards (and for the period 2025-2028 we linearly interpolate between the base rates and the 2024 rates). We represent the uncertainty in this multiplier using a gamma distribution with a mean of 1 and standard deviation of 0.58. This distribution has 2.5 and 97.5 percentiles of 0.20 and 2.42 respectively. The upper percentile thus corresponds to what might be expected in a worst-case scenario, where high levels of asymptomatic treatment initiation lead to lower levels of retention. (The upper limit of 2.42 corresponds to annual discontinuation rates of 1.01 and 0.78 in men and women respectively.) The lower limit, on the other hand, corresponds to a best-case scenario in which combinations of the previously described interventions lead to substantial reductions in treatment discontinuation.

Because the worst-case scenario is associated with higher rates of asymptomatic TB treatment association, we assume a negative correlation (r = -0.5) between the discontinuation rate multiplier and the time to door-to-door testing. (Although door-to-door testing is not the only strategy that would increase the future number of people with asymptomatic TB starting treatment, we expect it would be the most significant contributor.)

#### **5.4 Rate of cure in people completing treatment**

Many of the factors that affect rates of treatment discontinuation also affect rates of treatment adherence and cure. For example, community models of DOTS delivery may be more effective than the current model of clinic-based DOTS delivery. In a systematic review and meta-analysis, community and home-based DOTS were found to outperform clinic-based DOTS in terms of treatment success rates and sputum conversion rates (a more direct proxy for cure) [116]. The same review found that patient education interventions significantly improved treatment adherence (OR 1.83) and cure (OR 2.15), although each effect was estimated based on a single study. The review also found mostly positive evidence regarding the benefits of patient reminders in terms of adherence, sputum conversion and cure rates; for example, sputum conversion rates increased by an average OR of 1.29 (95% CI: 1.12-1.50) in three RCTs [116].

In a systematic review of nutritional support, some evidence was found of a positive effect on treatment adherence, although it was noted that interventions were very heterogeneous and hence no meta-analysis was performed [121]. However, few trials of nutritional interventions have directly assessed cure rates [112].

There is a risk that future cure rates could deteriorate if South Africa were to transition to a 4-month TB treatment regimen. In the randomized trial of the 4-month regimen currently endorsed by the WHO, the odds of unfavourable TB outcomes (mostly representing failure to achieve sustained TB cure) in the 4-month treatment arm was 1.87 times that in the 6-month treatment arm [108].

In the current Thembisa TB model, we have separate assumptions for the probability of cure in people who complete TB treatment (0.96) and people who do not complete treatment (0.65) [83]. To represent possible changes to future rates of cure, we apply an odds ratio adjustment to these defaults, which applies from 2029 onwards (with the rates between 2024 and 2029 changing linearly between the estimates for 2024 and 2029). To represent uncertainty regarding the value of this odds ratio, we specify a gamma prior distribution, with a mean of 1 and a standard deviation of 0.3. This means that the average future cure rate is the same as historic cure rates. The 2.5 and 97.5 percentiles of this gamma distribution are 0.50 and 1.67 respectively. The 0.50 corresponds to a worst-case scenario in which a new drug regimen is adopted with worse treatment failure rates (0.5 is similar to 0.53, the inverse of the odds of failure in the trial comparing 4-month and 6-month treatment regimens (1/1.87), i.e. approximating the odds of cure as the inverse of the odds of failure). The 1.67 corresponds to a best-case scenario in which one of the previously-cited interventions are implemented nationally and is effective in improving cure rates (1.7 is the average of the odds ratios for the effects of the previously cited patient education and reminder interventions on sputum conversion/cure).

Although we have linked the ‘worst-case’ scenario to the introduction of a shorter 4-month regimen, it is important to note that the net effect of such a change on cure rates can still be positive if patients are more likely to complete the 4-month regimen than they would the 6-month regimen. Consider the following hypothetical scenario:

- A 4-month regimen is introduced and the odds ratio for its effect on cure (controlling for treatment completion/default) is 0.75 when compared against the 6-month regimen. This means that under this regimen, the proportions cured are 0.947 and 0.582 in treatment completers and defaulters respectively.
- The proportion of people completing treatment increases from 88% (under the 6-month regimen) to 94% (under-the 4-month regimen).
- This means that the total proportion cured has increased slightly, from 92.3% under the 6-month regimen (0.88 × 0.96 + 0.12 × 0.65) to 92.5% under the 4-month regimen (0.94 × 0.947 + 0.06 × 0.582).

Because the assumptions about future cure rates and future treatment durations are somewhat linked, we assume a positive correlation (0.5) between the previously described odds ratio and the time to introducing a 4-month treatment regimen. This means we sample from a joint prior distribution for these two parameters rather than sampling independently from the two prior distributions.

38. Department of Health. **The South African National Tuberculosis Control Programme Practical Guidelines**. In; 2004.

39. Department of Health. **National Guidelines on the Treatment of Tuberculosis Infection**. In; 2023.

62. Massyn N, Day C, Barron P, Haynes R, English R, Padarath A. **District Health Barometer 2011/12**. In. Durban: Health Systems Trust; 2013.

63. Hermans S, Cornell M, Middelkoop K, Wood R. **The differential impact of HIV and antiretroviral therapy on gender-specific tuberculosis rates**. *Trop Med Int Health* 2019; 24(4):454-462.

64. Rees K, Muditambi N, Maswanganyi M, Railton J, McIntyre JA, Struthers HE, et al. **The impact of implementing a Xpert MTB/RIF algorithm on drug-sensitive pulmonary tuberculosis: a retrospective analysis**. *Epidemiology and Infection* 2018; 146(2):246-255.

65. Budgell EP, Evans D, Schnippel K, Ive P, Long L, Rosen S. **Outcomes of treatment of drug-susceptible tuberculosis at public sector primary healthcare clinics in Johannesburg, South Africa: A retrospective cohort study**. *S Afr Med J* 2016; 106(10):1002-1009.

66. Department of Health. **National HIV counselling and testing policy guidelines**. In; 2015.

74. World Health Organization. **WHO consolidated guidelines on tuberculosis. Module 2: Screening - systematic screening for tuberculosis disease**. In. Geneva; 2021.

75. van't Hoog AH, Langendam MW, Mitchell E, Cobelens FG, Sinclair D, Leeflang MMG, Lonnroth K. **A systematic review of the sensitivity and specificity of symptom- and chest-radiography screening for active pulmonary tuberculosis in HIV-negative persons and persons with unknown HIV status**. In; 2013.

83. Johnson LF, Kubjane M. **Thembisa TB, version 2.0: A model of tuberculosis in South Africa**. In. Cape Town: Centre for Infectious Disease Epidemiology and Research, University of Cape Town; 2024.

101. Naidoo P, Theron G, Rangaka MX, Chihota VN, Vaughan L, Brey ZO, Pillay Y. **The South African tuberculosis care cascade: estimated losses and methodological challenges**. *Journal of Infectious Diseases* 2017; 216 (Suppl 7):S702-S713.

110. World Health Organization. **WHO Consolidated Guidelines on Tuberculosis. Module 4: Treatment - Drug-susceptible TB Treatment**. In. Geneva; 2022.
